# Discharge prediction of critical patients with spinal cord injury: a machine learning study with 1485 cases

**DOI:** 10.1101/2021.06.26.21259569

**Authors:** Guoxin Fan, Huaqing Liu, Sheng Yang, Libo Luo, Lunji Wang, Mao Pang, Bin Liu, Liangming Zhang, Lanqing Han, Limin Rong

**Author notes:** Corresponding authors: Lanqing Han;, Limin Rong. These three authors equally contributed to this work (Guoxin Fan, Huaqing Liu, Sheng Yang).

## Abstract

**Objectives:** Prognostication of spinal cord injury (SCI) is vital, especially for critical patients who need intensive care. The study aims to develop machine-learning (ML) classifiers for discharge prediction of SCI patients in the intensive care unit (ICU).

**Methods:** Clinical data of patients diagnosed with SCI were extracted from the publicly available ICU database. A total of 105 ML classifiers were initially developed to predict the discharge destination (dead, further medical care, home), and then the top 3 classifiers with the best performance were stacked into an ensemble classifier (Esb-Clf). To balance the accuracy and the feasibility, the complete Esb-Clf was finally simplified with top 10 features (simplified Esb-Clf). The micro-average area under the curve (AUC) was used to compare the prediction performance of difference ML classifiers and 6 doctors’ artificial prediction.

**Results:** A total of 1485 SCI patients were used for the early and the recent prediction of discharge destination. In the early prediction, the micro-average AUC of the Esb-Clf and the simplified Esb-Clf was 0.846 and 0.835 during the independent testing, respectively. While in the recent prediction, the micro-average AUC of the Esb-Clf and the simplified Esb-Clf was 0.898 and 0.892. Performance of both the Esb-Clf and the simplified Esb-Clf were superior to the doctors’ in the early and the recent prediction.

**Conclusions:** ML classifiers can discriminate the discharge destination of SCI patients with high accuracy, feasibility and interpretability. Whether the simplified Esb-Clf as an online predictive tool is applicable to guiding clinical management needs further verification.

## 1. Introduction

It was reported that the annual incidence of spinal cord injury (SCI) in developed countries and regions ranged from 11.5 to 53.4/million[1]. With a prevalence of 583 per million[2], SCI results in a heavy economic burden to patients and society, with an estimated cost of $4 billion per year in the United States in terms of management of all SCI patients[1]. Additionally, the cost did not include the loss of income and the productivity of the patients, not to mention the spiritual loss and indirect health care expenditure. More importantly, critical SCI patients may suffer from a higher mortality due to renal failure, respiratory failure, hemodynamic instability, sepsis from urinary tract infection, life-threatening autonomic hyper-reflexia etc.[3-8]

Prognostication plays an important part in clinical practice, which can contribute to the decision-making of clinical management for doctors and assist the communication with the patients and their families. For critical patients in the intensive care unit (ICU), discharge destination may be one of the most important concerns[9], [10]. SCI patients with severe complications may die in the hospital, and those survived patients may need further medical care (FMC) like long-term professional nursing and rehabilitation[4]. Only a few survived SCI patients may go home after hospital discharge[4]. Thus, prognostication is vital for critical SCI patients. Although prognostication may be feasible at the group level, a patient-specific prediction could be challenging due to the heterogenous nature of the disease and significant variation within the cohort[11]. However, there is no personalized prediction model for individual prognosis of critical SCI patients in ICU.

Recently, the utility of machine learning (ML) in the fields of clinical and translational medicine is emerging[12-16]. The practicality of ML for prognostication has been widely validated[17-19], and many studies have adopted ML to predict hospital discharge[20-22]. While an early prediction at the time of admission may provide more time for clinicians to take actions on incoming situations[23], a recent prediction seems to be more accurate[24]. Therefore, the study aimed to develop ML models for early and recent prediction of discharge destination for critical SCI patients who received intensive care.

## 2. Methods

### 2.1 Database

All clinical data in the current study were extracted from the Medical Information Mart for Intensive Care III-IV (MIMIC-III-IV Database) and the eICU Collaborative Research Database (eICU Database). The MIMIC-III-IV database was established based on over 380000 patients from Beth Israel Deaconess Medical Center [25]. The MIMIC-III Database collected more than 58000 times hospitalization records of patients admitted to Beth Israel Deaconess Medical Center from June 2001 to October 2012. The MIMIC-IV Database collected clinical data from 2008 to 2019, so we combined these two databases (MIMIC-III and MIMIC-IV) as the MIMIC-III-IV Database from 2001 to 2019. The MIMIC-III-IV Database contains 26 tables in comma-separated values (CSV) format for inquiry. These tables include almost all data of patients during ICU treatment, such as diagnosis, laboratory examination data, demographic characteristics, transfer during hospitalization, treatment process, etc.

The eICU Database is a large and freely available multicenter database for critical care research supported by Philips Healthcare and the Laboratory for Computational Physiology at the Massachusetts Institute of Technology[26]. It comprises 200859 patient unit encounters for 139367 unique patients admitted between 2014 and 2015. The creation of the eICU Database is to expand the success of the MIMIC-III-IV Database to multiple centers. However, the source hospital of the MIMIC-III-IV Database is not involved in the eICU program, so the eICU Database is completely independent collected from numerous hospitals around the United States. The eICU Database contains 31 tables in CSV format for inquiry. These tables also include diagnosis, laboratory examination data, demographic characteristics, transfer during hospitalization, treatment process and so on.

### 2.2 Standard protocol approvals, registrations, and patient consents

This study protocol, which relies on two deidentified public database, was deemed exempt by the Institutional Review Board of our institution.

### 2.3 Data extraction

First, subject_id, hadm_id and icustay_id of patients diagnosed with SCI were obtained with the use of structured query language (SQL) language from the MIMIC-III-IV Database according to table diagnoses_icd, icustays and d_icd_diagnoses. Subject_id is the unique identifier of patients, that is, one subject_id corresponds to one patient, while one hadm_id represents one hospitalization record and icustay_id refers to the number given to patients when admitted to one specific ICU. Similar pieces of information were extracted from eICU database as uniquepid, patienthealthsystemstayid and patientunitstayid, respectively. We only included the first admission record for patients who had re-admissions more than once. After that, basic information of eligible patients was extracted from table admissions. In addition, laboratory examination data and physical sign data were acquired in the same way from table labevents and chartevents, looking up in definition-table d_labitems and d_items. Medication usage data was obtained from table prescriptions and inputevents. The time patient in and out of ICU was extracted from table icustays. The Glasgow scores of patients were also calculated. Similar extractions were adopted to the eICU Database.

Demographics information included ethnicity, gender, age, body mass index (BMI), etc. Vital signs included respiration rate, heart rate, systolic and diastolic pressure, mean arterial pressure. Laboratory data consisted of white blood cell count, red blood cell (RBC), platelet count, basophils, eosinophils, neutrophils, lymphocyte, monocyte, red blood cell distribution width (RDW), hemoglobin, hematocrit, mean corpuscular hemoglobin (MCH), mean corpuscular hemoglobin concentration (MCHC), mean corpuscular volume (MCV), prothrombin time (PT), activated partial thromboplastin time (APTT), international normalized ratio (INR), PaO2, PaCO2, FiO2, PH, bicarbonate, lactate, base excess (BE), anion gap, potassium, sodium, calcium, magnesium, chloride, phosphate, blood urea nitrogen (BUN), creatinine, albumin, blood glucose, etc. Medicine usage and treatment incorporated mechanical ventilation, morphine sulfate, cefazolin, potassium chloride (KCl), glucocorticoid, dopamine, dobutamine, epinephrine and norepinephrine. Missing data (<70%) were processed by different methods of imputation according to the variable type of features[27]. Predictive mean matching, logistic regression, polynomial regression was used for continuous, binary and categorical features respectively.

### 2.4 ML modeling

We extracted the clinical data of SCI patients from the MIMIC-III-IV Database and the eICU Database for discharge prediction, and then randomly divided the included data into sub-datasets (training: validation: testing= 6:2: 2). All training went through 3-time repeated 5-fold cross-validation with the training dataset, and all developed models underwent independent test with the testing dataset.

Seven common feature selection methods were conducted to identify useful variables for training ML models. The feature selection methods included maximal information coefficient (MIC), recursive feature elimination (RFE), embedding linear supported vector classifier (embedding LSVC), embedding logistic regressor (embedding LR), embedding tree, embedding random forest (RF), and minimal-redundancy-maximal-relevance (mRMR).

The selected features were then used to train fifteen ML algorithms, namely logistic regression, linear discriminant analysis (LDA), support vector machine (SVM), K-Nearest Neighbor (KNN), Gaussian Naïve Bayes (NB), decision tree, extra tree, random forest (RF), bagging, adaptive boosting (AdaBoost), gradient boosting decision tree (GBDT), light gradient boosting model (lightGBM), extreme gradient boosting (XGBoost), multilayer perceptron (MLP), and deep neural network (DNN). As a result, a total of 105 prediction classifiers from 7×15 combinations were initially developed for every prediction task.

### 2.5 Outcomes measures

An early prediction of the discharge destination (dead, FMC, home) was achieved with the first recorded data. Similarly, a recent prediction was conducted with the last recorded data. As lead time may contribute to the recent prediction for discharge destination, we added the lead time (e. g. length of stay in ICU (los-ICU), los-hospital, los before ICU admission, and los after ICU admission) as potential features for the recent prediction.

All predictions for ML models were category outcomes. The area under the curve (AUC) of the receiver operating characteristic (ROC) was the primary indicator to assess all ML classifiers. We selected the top 3 ML classifiers as they performed the best in averaged AUC in cross-validation. Then, we used a logistic regressor to further stack the top 3 ML classifiers into an ensemble classifier (Esb-Clf). To balance the accuracy and the feasibility, the Esb-Clf was finally simplified with 10 features (simplified Esb-Clf).

As for the rationale of the simplified Esb-Clf, we would present the variable AUC of the Esb-Clf along with different numbers of top features, which were rated by feature importance[28]. An approximate of 10 predictors would be selected, because most people were only able to interpret 7 to 10 features at a time[29]. Additionally, 6 doctors with different levels (A1-2 were senior doctors; B1-2 were attending doctors; C1-2 were resident doctors) were asked to conduct predictions with the same testing data, and their predictions were also compared with the complete Esb-Clf and the simplified Esb-Clf. To demonstrate their prediction performance, the micro-average AUC, sensitivity, and specificity were obtained for all mentioned classifiers. For the purpose of assisting clinical practice and academic research, we also developed the simplified Esb-Clf into an online predictive tool and disclose its code on the web.

### 2.6 Statistical analyses

Categorical variables were expressed as count (percentage), and the chi-square test or Kruskal-Wallis method was used for statistical tests. Continuous variables were described as mean (standard deviations), and group differences were tested by One-way ANOVA. All the data was analyzed with Python 3.7. Two-sided P values of < 0.05 were considered statistically significant.

### 2.7 Data Availability

The online predictive tool is presented at (https://colab.research.google.com/github/Huatsing-Lau/Prognosis_Prediction_Tool_of_SCI_Patients/blob/main/demo.ipynb), and its code is available from https://github.com/Huatsing-Lau/Prognosis_Prediction_Tool_of_SCI_Patients. Original data can be obtained from the MIMIC database (https://mimic.physionet.org/ for MIMIC-III and https://mimic-iv.mit.edu/ for MIMIC-IV) and the eICU database (https://eicu-crd.mit.edu/). Data not provided in the article because of space limitations may be shared (anonymized) at the request of any qualified investigator for purposes of replicating procedures and results.

## 3. Results

We initially identified 1599 ICU admissions records of 839 SCI patients from the MIMIC-III-IV database and 760 patients from the eICU database. After merging the two datasets and excluding patients with missing outcome (discharge destination), we finally included 1485 patients for prediction of discharge destination (**Fig 1**).

**Fig 1.**
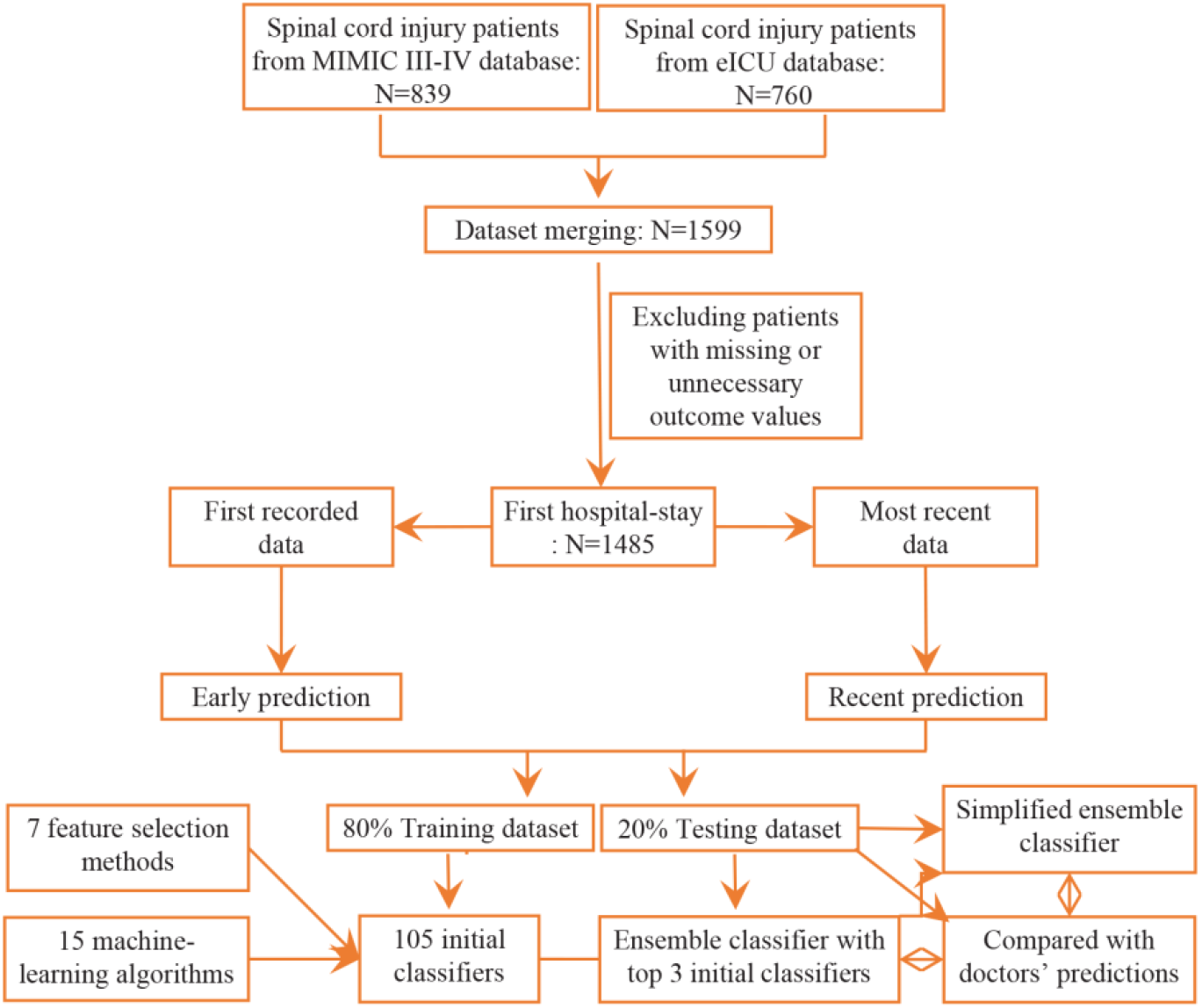
Flow chart of the development of prediction classifiers.

Basic characteristics of included patients for the early prediction and the recent prediction were shown in **Table 1** (Laboratory examination and treatment in **Table S1**). In total, 67 variables were included in our study. There were missing data in several variables including sum_diagnosis, ethnicity, age, gender, careunit, WBC, HR, respiration rate, diastolic arterial BP, systolic arterial BP, mean arterial BP, potassium, sodium, PO2, PT, PTT, blood glucose, hemoglobin, RDW, BUN, platelets, creatinine, bicarbonate, hematocrit, lactate and INR, etc. The percentage of missing values were shown in supplemental **Table S2-3**, range from 0 to 60.5%.

**Table 1.**
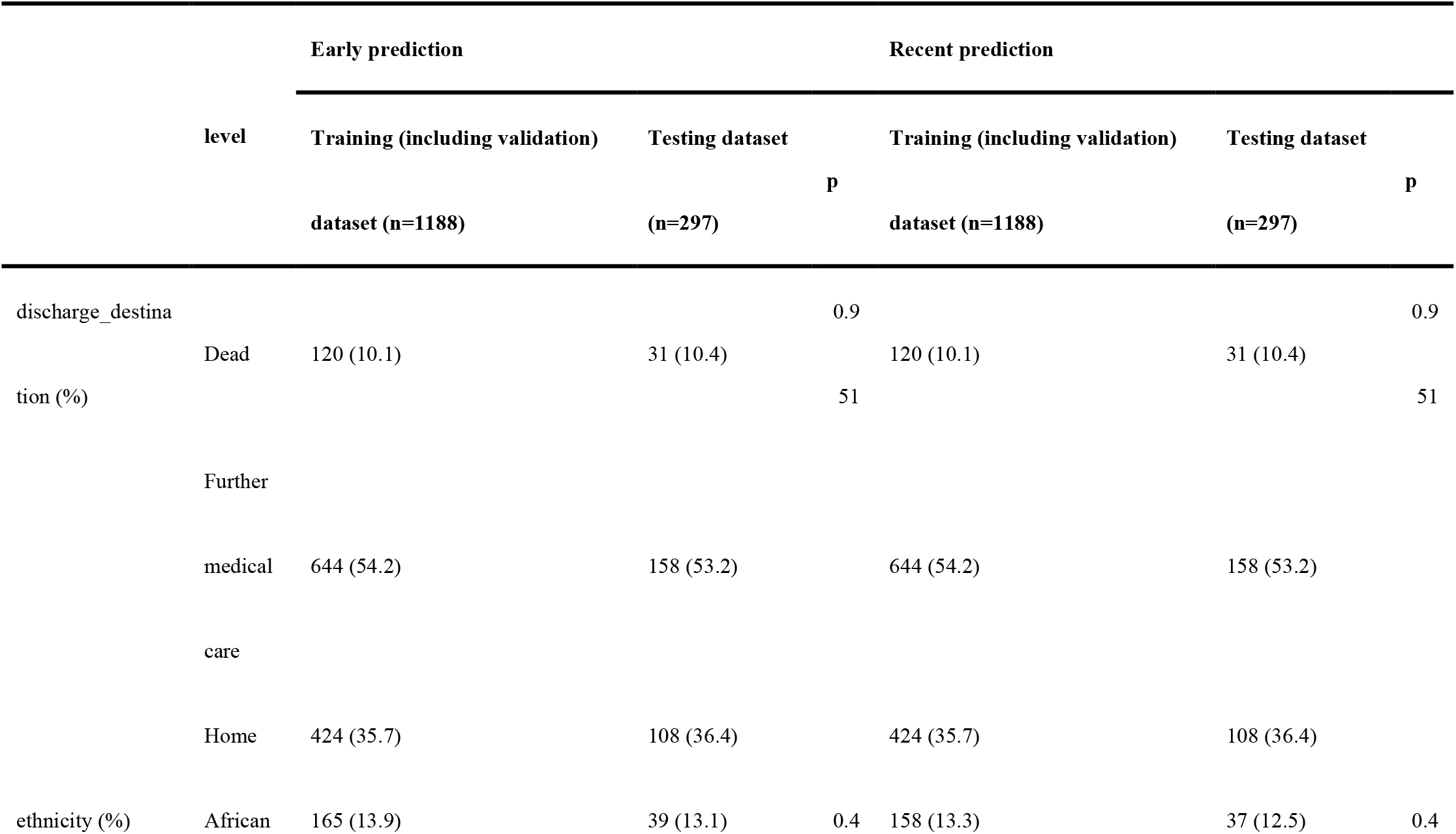
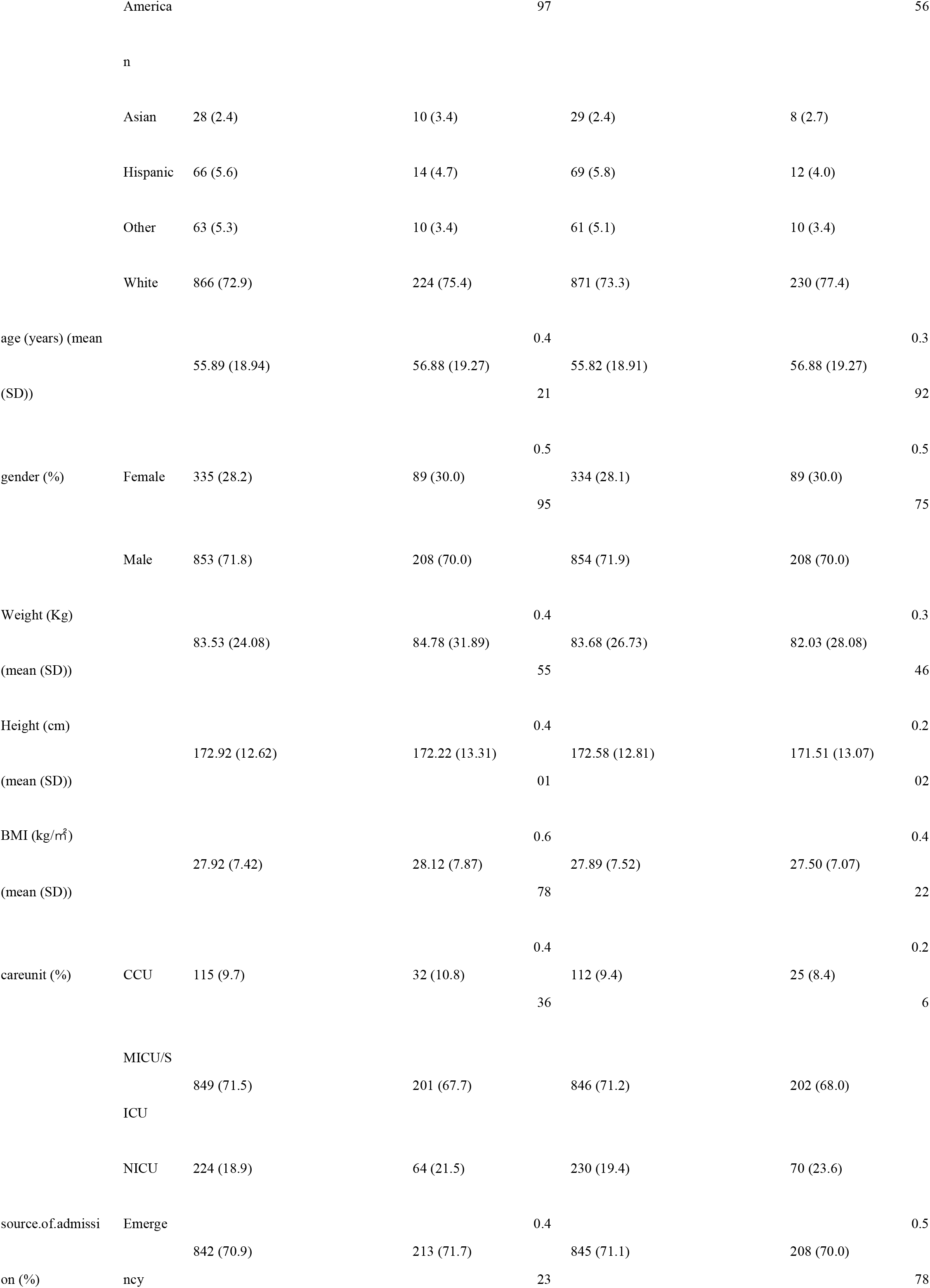
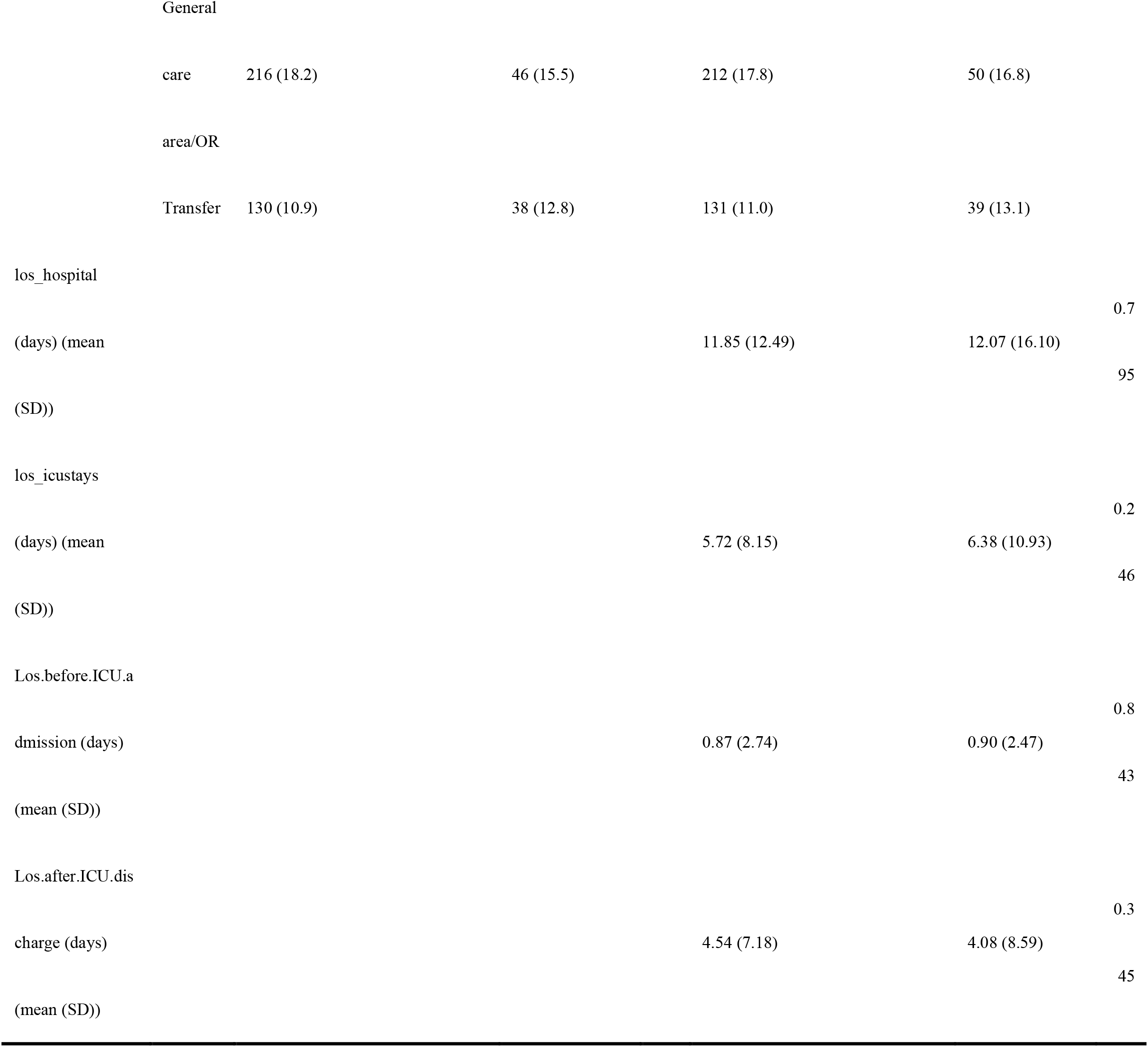
Basic characteristics of patients for discharge prediction.

For the early prediction, AUC for all the initial 105 classifiers were shown as a matrix (**Fig 2A, B**). Comparing the micro-average AUC during cross-validation, the top 3 initial ML classifiers for the early prediction were SVM (embedding tree), bagging (embedding tree) and bagging (embedding LSVC). After stacking, the AUC of complete Esb-Clf (micro-average AUC: 0.846) was slightly higher than the average AUC of the top 3 ML classifiers during independent testing (micro-average AUC: 0.832, 0.835, 0.84). The ROC of the simplified Esb-Clf was quite similar to the complete Esb-Clf in the early prediction (**Fig 3**), and the micro-average AUC (0.835) of simplified Esb-Clf was comparable to the complete Esb-Clf, both of which were superior to doctors with different levels (micro-average AUC: 0.57-0.74).

**Fig 2.**
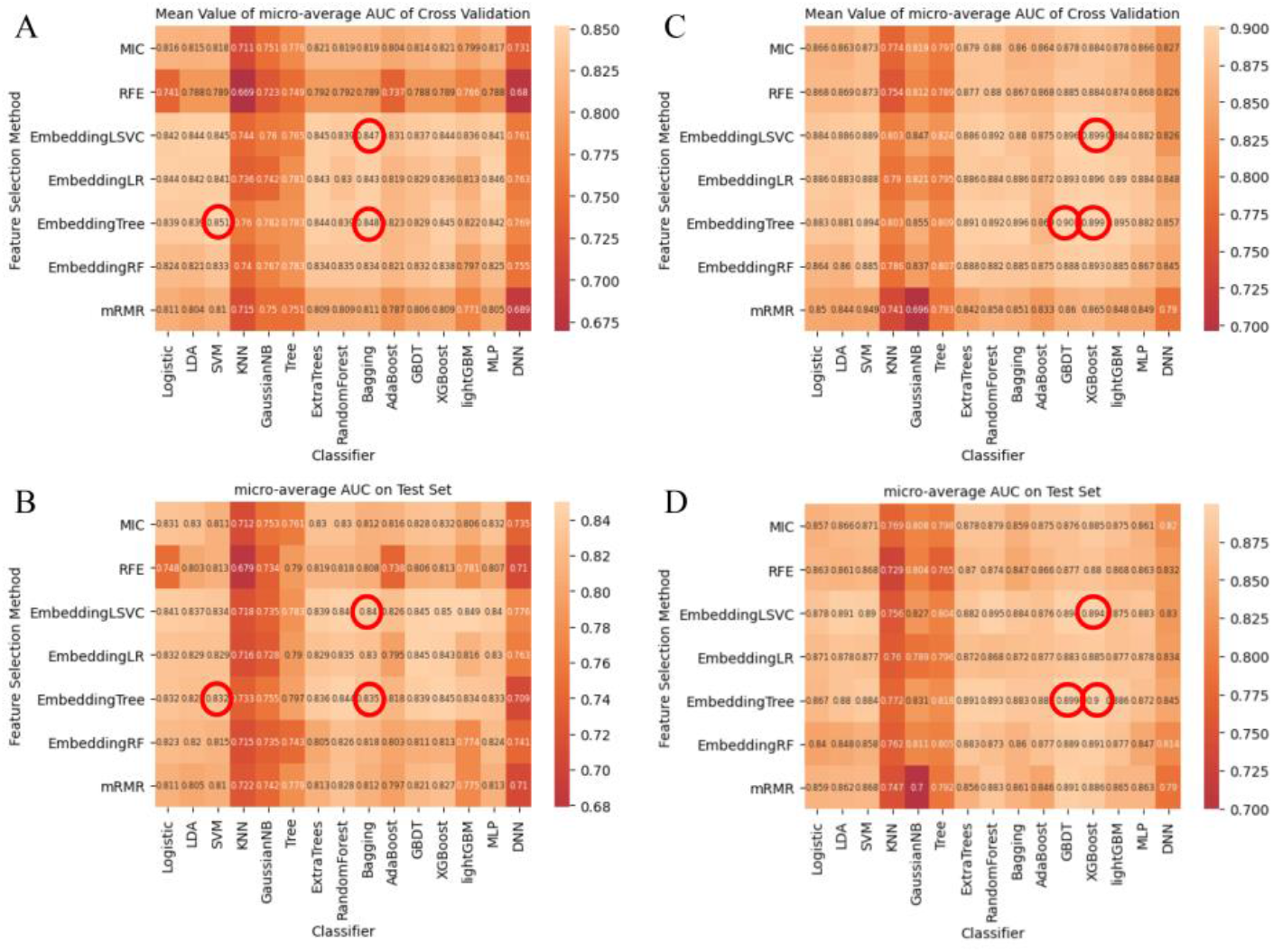
AUC of the initial 105 classifiers for discharge prediction. A: cross-validation for the early prediction; B: independent testing for the early prediction; C: cross-validation for the recent prediction; D: independent testing for the recent prediction.

**Fig 3.**
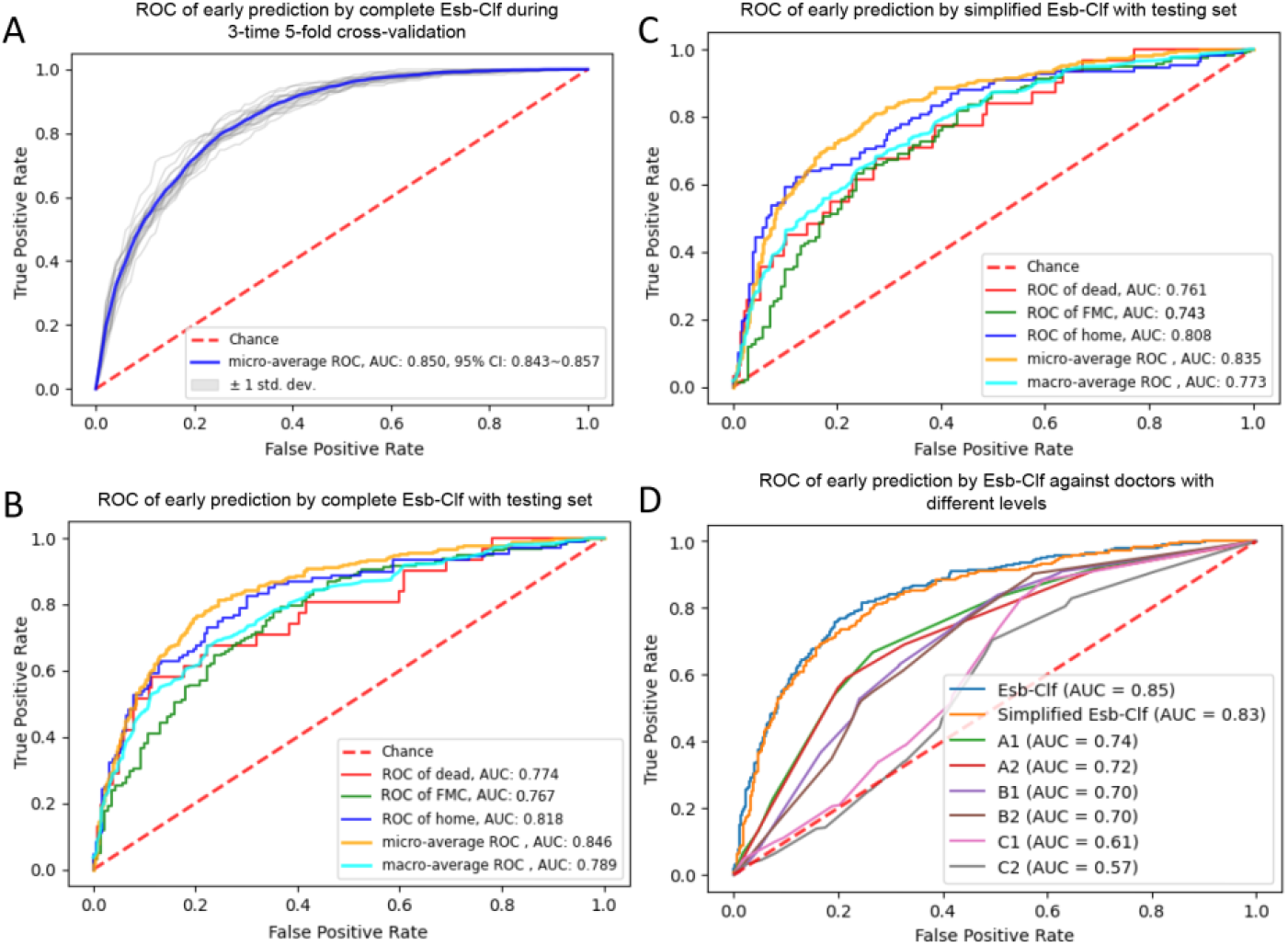
ROC of the complete Esb-Clf and the simplified Esb-Clf for the early prediction. A: the performance of the complete Esb-Clf during cross-validation; B: the performance of the complete Esb-Clf during independent testing; C: the performance of the simplified Esb-Clf during independent testing; D: the performance of the complete Esb-Clf and the simplified Esb-Clf compared with different-level doctors.

For the recent prediction, AUC for all the initial 105 classifiers were shown as a matrix (**Fig 2C, D**). Comparing the micro-average AUC during cross-validation, the top 3 initial ML classifiers for the recent prediction were GBDT (embedding tree), XGBoost (embedding tree) and XGBoost (embedding LSVC). After stacking, the AUC of complete Esb-Clf (micro-average AUC: 0.898) was slightly higher than the average AUC of the top 3 ML classifiers during independent testing (micro-average AUC: 0.894, 0.899, 0.9). The ROC of the simplified Esb-Clf was quite similar to the complete Esb-Clf in the recent prediction (**Fig 4**), and the micro-average AUC (0.892) of simplified Esb-Clf was comparable to the complete Esb-Clf, both of which were superior to doctors with different levels (micro-average AUC: 0.62-0.71).

**Fig 4.**
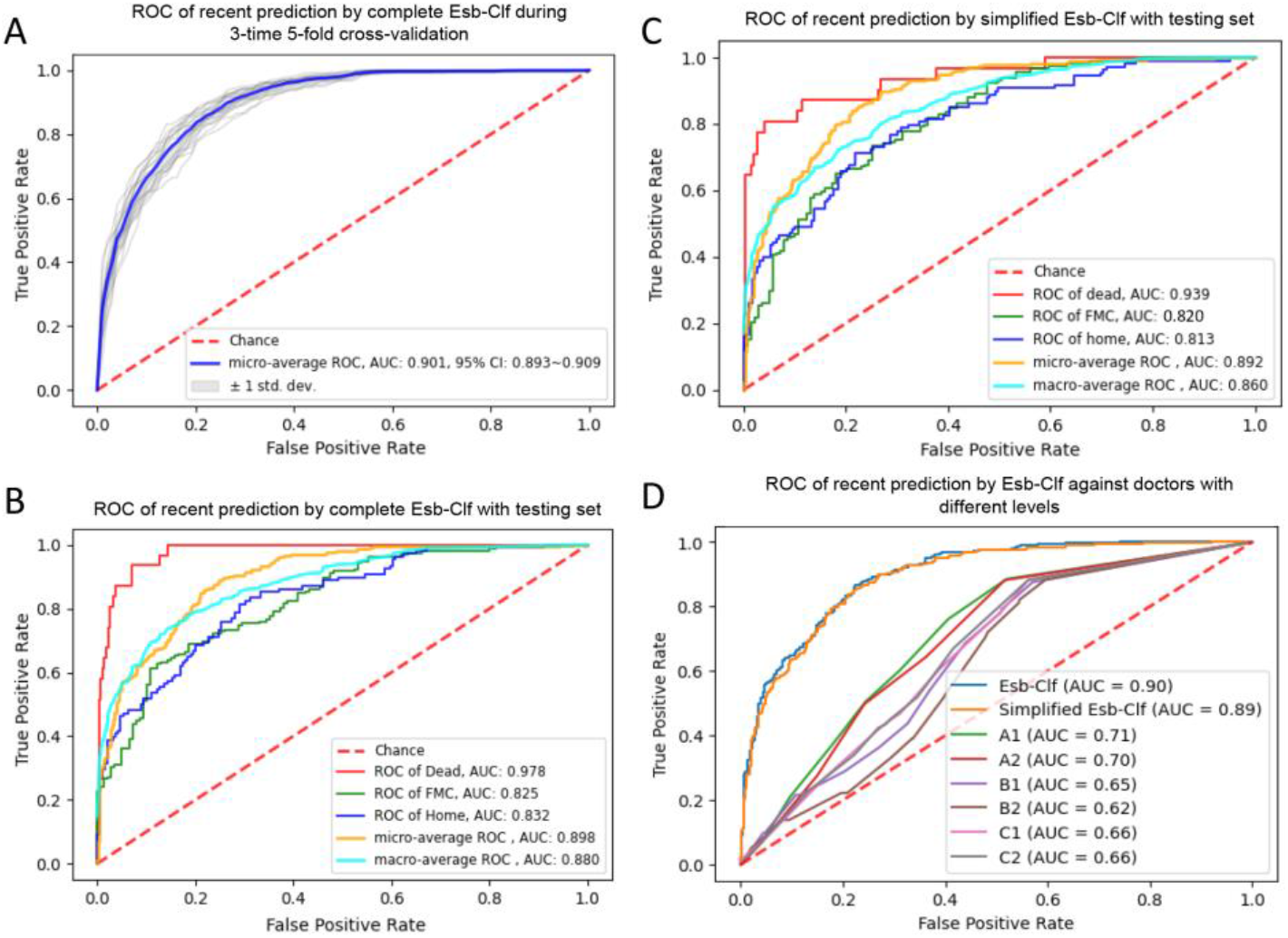
ROC of the complete Esb-Clf and the simplified Esb-Clf for the recent prediction. A: the performance of the complete Esb-Clf during cross-validation; B: the performance of the complete Esb-Clf during independent testing; C: the performance of the simplified Esb-Clf during independent testing; D: the performance of the complete Esb-Clf and the simplified Esb-Clf compared with different-level doctors.

As for comparisons of specific prediction targets (i.e. dead, FMC, home), the simplified Esb-Clf and the complete Esb-Clf defeated the doctors mainly in early prediction of FMC (P values < 0.05 in all Delong tests). Delong tests of the early predictions for all classifiers could be found in supplemental **Fig S1-3**. The AUC values, sensitivity, and specificity of all classifiers for the early prediction of specific prediction targets were also shown in **Fig 5A**. Similarly, the simplified Esb-Clf and the complete Esb-Clf defeated the doctors mainly in recent prediction of FMC and home (P values < 0.05 in all Delong tests). Delong tests of the recent predictions for all classifiers could be found in supplemental **Fig S4-6**. The AUC values, sensitivity, and specificity of all classifiers for the recent prediction of specific prediction targets were also shown in **Fig 5B**. Considering the feasibility over the complete Esb-Clf and the accuracy superiority over the doctors, we developed the simplified Esb-Clf into an online prediction tool to achieve early and recent prediction of discharge destination for critical SCI patients (**Fig 5C**).

**Fig 5.**
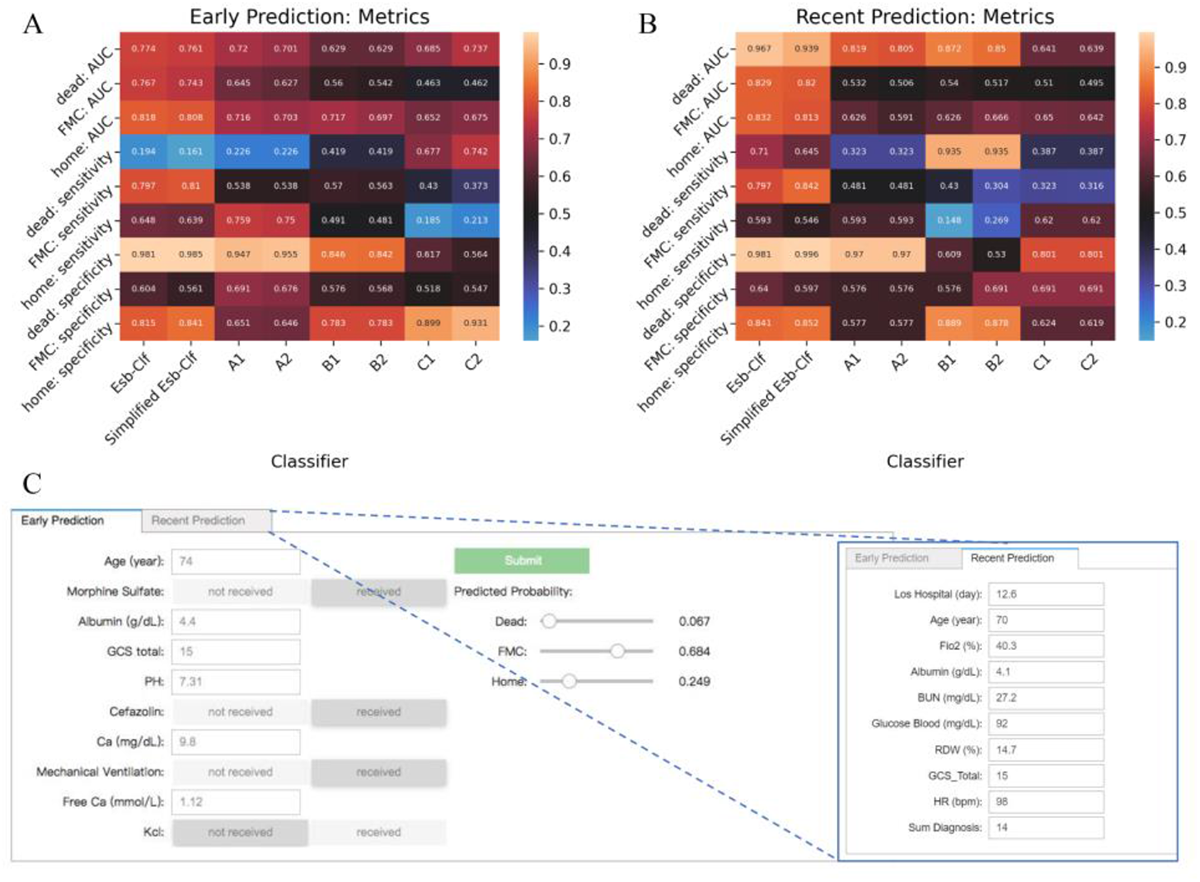
A: AUC values, sensitivity, and specificity of all classifiers for the early prediction; B: AUC values, sensitivity, and specificity of all classifiers for the recent prediction; C: online prediction tool based on the simplified Esb-Clf.

The variable AUC of the Esb-Clf along with different numbers of top features was shown in **Fig 6**. When top features were over 10, the AUC of the Esb-Clf was not significantly improved neither for the early prediction or the recent prediction. Thus, the simplified Esb-Clf was developed with top 10 features to balance the accuracy and the feasibility. The top 10 features of the simplified Esb-Clf for the early prediction were ‘age’, ‘morphine sulfate (received or not)’, ‘albumin’, ‘GCS total’, ‘PH’, ‘cefazolin (received or not)’, ‘Ca2+’, ‘mechanical ventilation (received or not)’, ‘free Ca2+’, and ‘KCl (received or not)’, namely. The top 10 features of the simplified Esb-Clf for the recent prediction were ‘los_hospital’, ‘age’, ‘FiO2’, ‘albumin, ‘BUN’, ‘glucose blood’, ‘RDW’, ‘GCS total’, ‘HR’, and ‘sum_diagnosis’, namely.

**Fig 6.**
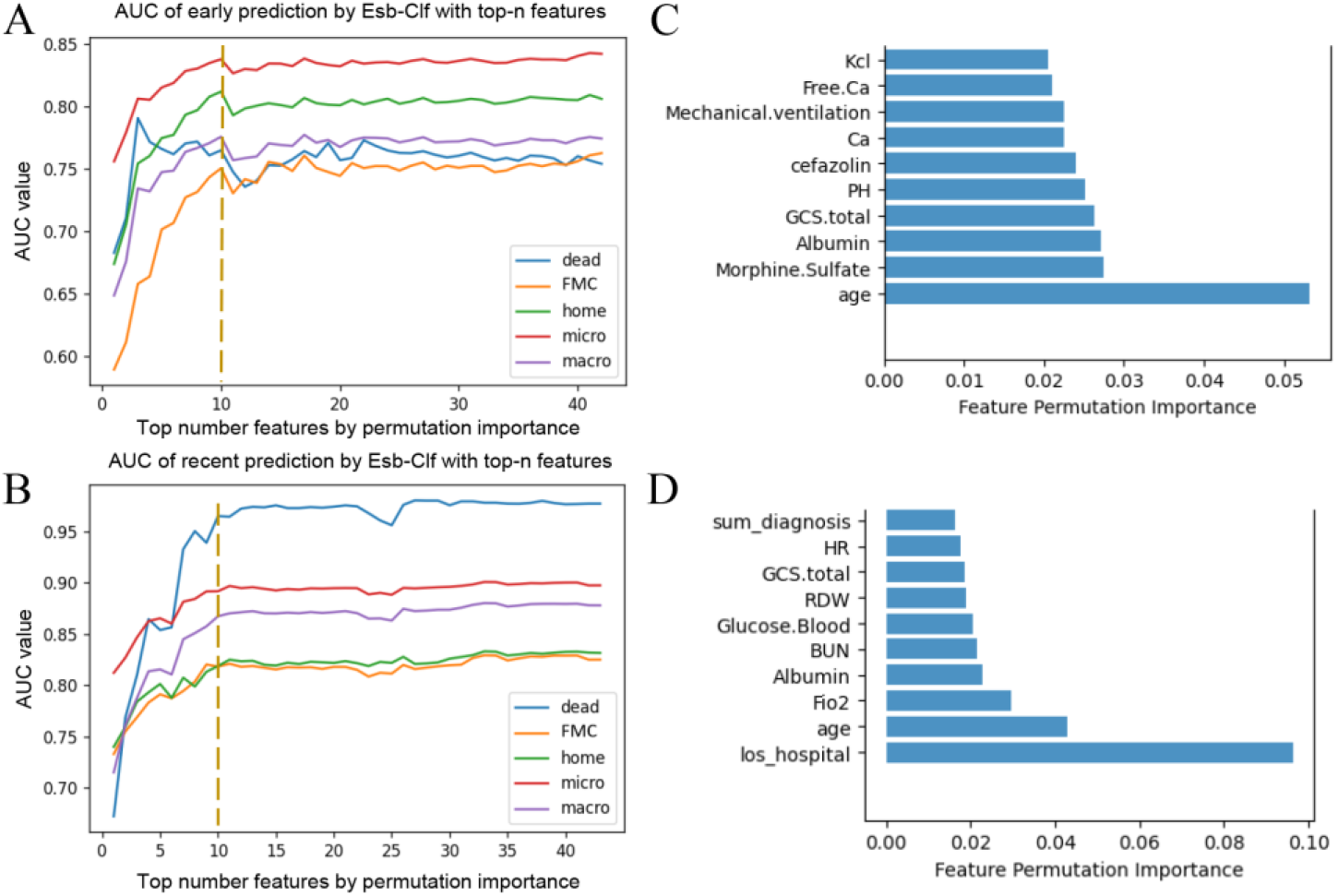
Variable AUC of the Esb-Clf and the interpretability of the simplified Esb-Clf. A: AUC of the Esb-Clf varied with features for the early prediction; B: AUC of the Esb-Clf varied with features for the recent prediction; C: top 10 features of the simplified Esb-Clf for the early prediction; D: top 10 features of the simplified Esb-Clf for the recent prediction.

## 4. Discussion

The prognosis of critical SCI patients who received intensive care could be very frustrated. Accurate prognostication might benefit the decision-making of the management. This study demonstrated that the simplified Esb-Clf accurately predicted the discharge destination of critical SCI patients both in the early admission and the later hospital stay, which was also associated with high feasibility and interpretability. The online predictive tool based on the simplified Esb-Clf might benefit the patients, their families, doctors and researchers in the future.

There is a paucity of research on the clinical outcomes of SCI patients who require ICU care [4], [8], [30-32]. The mortality rate of critical SCI patients was about 10% in the current study, but other reported the mortality rate could reach 38.5%[33]. For those survived SCI patients in ICU, most of them may need long-term rehabilitation and nursery support due to motor deficit, urinary dysfunction or defecation obstacle[30]. The percentage of critical SCI patients who need professional rehabilitation and nursery support (discharge to FMC) was about 50%, which was similar (about 45%) in the other study[4]. Nearly one third critical SCI patients were discharged home, which was higher than that (about 15%) in the other study[4]. Anyway, the imbalanced three-class categories did not significantly compromise the prediction accuracy in the current study. A micro-average AUC of 0.892-0.898 in the recent prediction was very promising, and a micro-average AUC of 0.835-0.846 in the early prediction was also acceptable.

ML was popular in predicting prognosis, as it prevailed at modeling non-linear relationships between the given predictors and the predictive outcomes[18]. Several studies[34-38] have developed ML models to predict the prognosis of SCI patients. With electroencephalography data, Vuckovic et al.[36] implemented ML to predict the central neuropathic pain in SCI patients with an accuracy of 0.83-0.86. With tabular data, DeVries Z et al.[39] proposed an unsupervised ML model to predict the walking ability of SCI patients with an overall accuracy of 0.72-0.90. Similarly, the current study also achieved high accuracy (AUC of 0.835-0.898) in predicting discharge destination of critical SCI patients. However, the current study adopted different feature selection methods and numerous ML algorithms, which extensively explored the potential of different combinations and improved the generalization. Furthermore, the prediction performance of the simplified Esb-Clf was superior to the doctors with different levels, who unsurprisingly found to be weak at integrating tabular information and allocating mild probabilities at three-class predictions. It seemed that the incompetent predictions of the doctors were mainly compromised by the classification of FMC and home.

Appropriate feature selection is necessary [40-42], because fewer features may compromise the predicting performance, while superabundant predictors may lead to over-fitting[43]. More importantly, feature selection enables a promising model to balance the accuracy, interpretability and feasibility. In the current study, the prediction accuracy of the initial 105 classifiers varied a lot among different feature selection methods, which was similar to other studies[44], [45]. However, it seemed that embedding LSVC and embedding tree were the top two feature selection methods, as they comprised the top 3 ML classifiers. After stacking the top 3 ML classifiers, only a slight improvement of AUC was observed for the complete Esb-Clf, but its generalization ability was definitely improved. Additionally, the generalization ability of the Esb-Clf was also guaranteed by multiple data sources, as it was trained with the multicenter ICU data all over the United States. After the simplification with the top 10 features, the accuracy of the simplified Esb-Clf was not compromised at all, but its feasibility potential as an online predictive tool was significantly improved.

While it is vital to pursue early prediction, a recent and accurate prediction may enable doctors to take prompt actions to deal with the critical situation. Based on the most recent clinical data, Temple et al.[20] used ML to identify which patients in neonatal intensive care could be discharged home in the next 2–10 days. In the current study, we found higher accuracy of discharge destination in the recent prediction compared with that in the early prediction. This might not be a surprising finding, because predictive features extracted from the last examination were more closely correlated to patient conditions. However, the top 10 features were quite different in the early and the recent prediction. Only ‘age’, ‘albumin’ and ‘GCS.total’ were common in the two predictions. Interestingly, lead time like ‘los_hospital’ did contribute a lot to the recent prediction, which was similar to other studies[10], [20]. Unexpectedly, this feature even rated top 1 importance in the recent prediction, which might be hard to be rated and integrated into the prediction by doctors. Anyway, the interpretability of the simplified Esb-Clf certainly improved the feasibility further in practice.

Nevertheless, the current research also has limitations. First, the dataset did not record the information of grading or scoring the injury degree, which might contribute to the prediction performance of ML classifiers. Second, the current study could not realize prediction of long-term outcomes of SCI patients who underwent intensive medical care, because the eICU database did not include the follow-up information. Third, the online predictive tool might assist the doctors in informing the patients or their families of the discharge destination. However, whether the online predictive tool is applicable to guiding clinical treatment or resource allocations needs further verification. We hope more researchers get interested in this topic and further explore the clinical potential of the online predictive tool.

## 5. Conclusions

The simplified Esb-Clf accurately discriminate the discharge destination in the recent prediction, and its performance in the early prediction was also acceptable, which indicated its potential as an online predictive tool with high accuracy, feasibility and interpretability. Whether the simplified Esb-Clf as an online predictive tool is applicable to guiding clinical management needs further verification.

## Supporting information

STROBE Checklist

Tripod Checklist

coi_disclosure

## Data Availability

The online predictive tool is presented at (https://colab.research.google.com/github/Huatsing-Lau/
Prognosis_Prediction_Tool_of_SCI_Patients/blob/main/demo.ipynb), and its code is available from https://github.com/Huatsing-Lau/Prognosis_Prediction_Tool_of_SCI_Patients. Original data can be obtained from the MIMIC database (https://mimic.physionet.org/ for MIMIC-III and https://mimic-iv.mit.edu/ for MIMIC-IV) and the eICU database (https://eicu-crd.mit.edu/). Data not provided in the article because of space limitations may be shared (anonymized) at the request of any qualified investigator for purposes of replicating procedures and results.

## Abbreviations

SCI: spinal cord injury
ML: machine-learning
ICU: intensive care unit
Esb-Clf: ensemble classifier
AUC: area under the curve
FMC: further medical care
CSV: comma-separated values
SQL: structured query language
BMI: body mass index
RBC: red blood cell
RDW: red blood cell distribution width
MCH: mean corpuscular hemoglobin
MCHC: mean corpuscular hemoglobin concentration
MCV: mean corpuscular volume
PT: prothrombin time
APTT: activated partial thromboplastin time
INR: international normalized ratio
BE: base excess
BUN: blood urea nitrogen
MIC: maximal information coefficient
RFE: recursive feature elimination
LSVC: linear supported vector classifier
LR: logistic regressor
RF: random forest
mRMR: minimal-redundancy-maximal-relevance
LDA: linear discriminant analysis
SVM: support vector machine
KNN: K-Nearest Neighbor
NB: Gaussian Naïve Bayes
AdaBoost: adaptive boosting
GBDT: gradient boosting decision tree
lightGBM: light gradient boosting model
XGBoost: extreme gradient boosting
MLP: multilayer perceptron
DNN: deep neural network
los: length of stay
ROC: receiver operating characteristic

