## Supplemental materials for "Discharge prediction of critical patients with spinal cord injury: a machine learning study with 1485 cases"

**Supplementary Table 1. Laboratory examination and treatment of patients for discharge prediction.**

|  | **level** | **Early prediction** | | | **Recent prediction** | | |
| --- | --- | --- | --- | --- | --- | --- | --- |
|  |  | **Training (including validation) dataset (n=1188)** | **Testing dataset (n=297)** | **p** | **Training (including validation) dataset (n=1188)** | **Testing dataset (n=297)** | **p** |
| sum_diagnosis (mean (SD)) |  | 11.56 (9.05) | 10.61 (8.41) | 0.1 | 11.53 (9.03) | 10.62 (8.40) | 0.118 |
| Paralysis (%) | No | 760 (64.0) | 187 (63.0) | 0.798 | 765 (64.4) | 181 (60.9) | 0.299 |
|  | Yes | 428 (36.0) | 110 (37.0) |  | 423 (35.6) | 116 (39.1) |  |
| Mechanical.ventilation (%) | No | 703 (59.2) | 166 (55.9) | 0.336 | 700 (58.9) | 161 (54.2) | 0.16 |
|  | Yes | 485 (40.8) | 131 (44.1) |  | 488 (41.1) | 136 (45.8) |  |
| GCS.total (mean (SD)) |  | 12.20 (4.23) | 12.42 (3.97) | 0.402 | 12.96 (3.74) | 13.23 (3.51) | 0.256 |
| HR (bpm) (mean (SD)) |  | 84.77 (21.24) | 84.10 (19.93) | 0.623 | 77.12 (25.69) | 73.51 (28.72) | 0.035 |
| respiration (bpm) (mean (SD)) |  | 17.72 (7.47) | 17.65 (8.09) | 0.887 | 17.05 (8.47) | 16.37 (8.75) | 0.221 |
| Arterial.BP..Diastolic. (mmHg) (mean (SD)) |  | 69.06 (17.96) | 71.78 (20.51) | 0.024 | 65.83 (17.79) | 65.12 (19.09) | 0.545 |
| Arterial.BP..Systolic. (mmHg) (mean (SD)) |  | 127.34 (26.90) | 128.43 (28.44) | 0.535 | 123.18 (27.17) | 119.29 (31.35) | 0.033 |
| Arterial.BP..mean. (mmHg) (mean (SD)) |  | 84.97 (20.21) | 87.49 (21.71) | 0.058 | 81.04 (20.00) | 79.73 (21.70) | 0.322 |
| Potassium (mmol/L) (mean (SD)) |  | 4.09 (0.66) | 4.02 (0.58) | 0.108 | 4.10 (0.50) | 4.11 (0.48) | 0.68 |
| Sodium (mmol/L) (mean (SD)) |  | 138.24 (4.46) | 138.41 (5.01) | 0.584 | 138.59 (4.24) | 138.57 (3.89) | 0.929 |
| PO2 (mmHg) (mean (SD)) |  | 155.06 (102.40) | 166.36 (104.65) | 0.091 | 121.26 (67.60) | 123.36 (66.76) | 0.633 |
| PT (seconds) (mean (SD)) |  | 14.16 (5.29) | 14.24 (6.02) | 0.821 | 14.13 (5.52) | 14.05 (4.62) | 0.81 |
| PTT (seconds) (mean (SD)) |  | 30.72 (13.75) | 31.16 (15.97) | 0.63 | 31.76 (12.40) | 31.00 (13.25) | 0.348 |
| Glucose.Blood (mg/dL) (mean (SD)) |  | 138.54 (105.10) | 133.83 (52.54) | 0.454 | 125.15 (44.86) | 123.40 (43.71) | 0.546 |
| Hemoglobin (g/dL) (mean (SD)) |  | 12.04 (2.32) | 12.26 (2.33) | 0.142 | 10.69 (1.95) | 10.90 (2.07) | 0.106 |
| RDW (%) (mean (SD)) |  | 14.28 (2.13) | 14.44 (2.00) | 0.253 | 14.80 (2.23) | 14.84 (2.26) | 0.756 |
| BUN (mg/dL) (mean (SD)) |  | 19.52 (14.30) | 19.86 (16.08) | 0.721 | 19.03 (15.20) | 18.44 (12.65) | 0.543 |
| WBC (K/uL) (mean (SD)) |  | 11.64 (6.26) | 11.58 (6.91) | 0.883 | 9.71 (4.83) | 10.33 (5.98) | 0.062 |
| Platelets (K/uL) (mean (SD)) |  | 235.23 (105.96) | 237.20 (120.26) | 0.781 | 268.38 (140.57) | 270.68 (141.16) | 0.801 |
| creatinine (mg/dL) (mean (SD)) |  | 1.05 (0.89) | 1.01 (0.73) | 0.567 | 0.86 (0.72) | 0.83 (0.59) | 0.506 |
| bicarbonate (mmol/L) (mean (SD)) |  | 24.06 (4.34) | 24.34 (4.13) | 0.31 | 26.68 (4.00) | 26.77 (3.82) | 0.729 |
| hematocrit (%) (mean (SD)) |  | 36.00 (6.53) | 36.80 (6.39) | 0.061 | 32.35 (5.43) | 33.02 (5.87) | 0.063 |
| lactate (mmol/L) (mean (SD)) |  | 2.02 (1.52) | 1.94 (1.37) | 0.401 | 1.65 (1.52) | 1.57 (1.39) | 0.374 |
| INR (mean (SD)) |  | 1.26 (0.58) | 1.27 (0.70) | 0.956 | 1.25 (0.56) | 1.23 (0.43) | 0.678 |
| BE (mmol/L) (mean (SD)) |  | -0.82 (5.11) | -0.72 (5.66) | 0.761 | 1.26 (4.74) | 1.48 (4.86) | 0.472 |
| Basophils (%) (mean (SD)) |  | 0.32 (0.38) | 0.37 (0.39) | 0.02 | 0.37 (0.43) | 0.32 (0.40) | 0.077 |
| Ca (mg/dL) (mean (SD)) |  | 8.53 (0.82) | 8.67 (0.75) | 0.008 | 8.61 (0.70) | 8.66 (0.63) | 0.265 |
| Chloride (mmol/L) (mean (SD)) |  | 103.59 (5.53) | 103.34 (5.58) | 0.49 | 102.84 (5.13) | 102.92 (4.84) | 0.797 |
| Free.Ca (mmol/L) (mean (SD)) |  | 0.95 (0.45) | 0.92 (0.45) | 0.251 | 0.97 (0.37) | 0.93 (0.39) | 0.145 |
| Phosphate (mg/dL) (mean (SD)) |  | 3.35 (1.06) | 3.34 (1.15) | 0.783 | 3.43 (1.03) | 3.39 (0.97) | 0.504 |
| Eosinophils (%) (mean (SD)) |  | 1.34 (1.81) | 1.42 (1.70) | 0.48 | 2.06 (2.37) | 1.58 (1.97) | 0.001 |
| Lymphocytes (%) (mean (SD)) |  | 14.89 (10.60) | 15.63 (11.30) | 0.292 | 16.19 (10.81) | 15.12 (10.36) | 0.125 |
| Magnesium (mg/dL) (mean (SD)) |  | 1.83 (0.33) | 1.86 (0.34) | 0.139 | 2.01 (0.29) | 1.99 (0.29) | 0.364 |
| Monocytes (%) (mean (SD)) |  | 6.25 (3.32) | 6.27 (3.00) | 0.917 | 6.88 (3.32) | 6.90 (5.59) | 0.963 |
| PCO2 (mmHg) (mean (SD)) |  | 42.66 (11.18) | 42.74 (11.43) | 0.909 | 41.33 (9.01) | 41.91 (10.80) | 0.339 |
| RBC (m/uL) (mean (SD)) |  | 4.02 (0.76) | 4.07 (0.77) | 0.264 | 3.60 (0.65) | 3.68 (0.70) | 0.062 |
| MCH (pg) (mean (SD)) |  | 29.89 (3.24) | 30.07 (2.58) | 0.375 | 29.83 (2.50) | 29.83 (2.39) | 0.983 |
| MCHC (%) (mean (SD)) |  | 33.31 (1.83) | 33.32 (1.43) | 0.975 | 33.01 (1.52) | 32.91 (1.43) | 0.312 |
| MCV (fL) (mean (SD)) |  | 89.65 (8.54) | 90.39 (6.74) | 0.165 | 90.37 (6.34) | 90.61 (6.34) | 0.561 |
| PH (mean (SD)) |  | 7.37 (0.10) | 7.37 (0.10) | 0.78 | 7.41 (0.07) | 7.41 (0.08) | 0.771 |
| Anion.Gap (mmol/L) (mean (SD)) |  | 13.00 (4.19) | 13.00 (4.29) | 0.993 | 11.43 (3.99) | 11.07 (4.15) | 0.175 |
| CO2.Blood (mmol/L) (mean (SD)) |  | 26.11 (6.53) | 26.44 (6.96) | 0.441 | 28.01 (7.29) | 28.70 (7.02) | 0.14 |
| Specific.Gravity (mean (SD)) |  | 1.02 (0.01) | 1.02 (0.01) | 0.931 | 1.02 (0.01) | 1.02 (0.01) | 0.624 |
| Fio2 (%) (mean (SD)) |  | 55.78 (29.77) | 55.83 (31.64) | 0.981 | 43.98 (22.41) | 43.10 (23.82) | 0.548 |
| Total.bilirubin (mg/dL) (mean (SD)) |  | 0.71 (0.88) | 0.75 (1.17) | 0.561 | 0.80 (1.86) | 0.75 (1.47) | 0.677 |
| Albumin (g/dL) (mean (SD)) |  | 3.21 (0.71) | 3.31 (0.69) | 0.046 | 3.15 (0.74) | 3.18 (0.72) | 0.56 |
| Morphine.Sulfate (%) | No | 798 (67.2) | 219 (73.7) | 0.035 | 801 (67.4) | 219 (73.7) | 0.043 |
|  | Yes | 390 (32.8) | 78 (26.3) |  | 387 (32.6) | 78 (26.3) |  |
| cefazolin (%) | No | 851 (71.6) | 220 (74.1) | 0.443 | 859 (72.3) | 220 (74.1) | 0.59 |
|  | Yes | 337 (28.4) | 77 (25.9) |  | 329 (27.7) | 77 (25.9) |  |
| Kcl (%) | No | 566 (47.6) | 151 (50.8) | 0.357 | 568 (47.8) | 151 (50.8) | 0.384 |
|  | Yes | 622 (52.4) | 146 (49.2) |  | 620 (52.2) | 146 (49.2) |  |
| glucocorticoid (%) | No | 887 (74.7) | 221 (74.4) | 0.988 | 894 (75.3) | 221 (74.4) | 0.822 |
|  | Yes | 301 (25.3) | 76 (25.6) |  | 294 (24.7) | 76 (25.6) |  |
| Dopamine (%) | No | 1149 (96.7) | 291 (98.0) | 0.344 | 1151 (96.9) | 290 (97.6) | 0.619 |
|  | Yes | 39 (3.3) | 6 (2.0) |  | 37 (3.1) | 7 (2.4) |  |
| Dobutamine (%) | No | 1181 (99.4) | 296 (99.7) | 0.929 | 1181 (99.4) | 296 (99.7) | 0.929 |
|  | Yes | 7 (0.6) | 1 (0.3) |  | 7 (0.6) | 1 (0.3) |  |
| Epinephrine (%) | No | 1156 (97.3) | 284 (95.6) | 0.185 | 1154 (97.1) | 284 (95.6) | 0.251 |
|  | Yes | 32 (2.7) | 13 (4.4) |  | 34 (2.9) | 13 (4.4) |  |
| Norepinephrine (%) | No | 990 (83.3) | 254 (85.5) | 0.408 | 998 (84.0) | 253 (85.2) | 0.682 |
|  | Yes | 198 (16.7) | 43 (14.5) |  | 190 (16.0) | 44 (14.8) |  |

**Supplementary Table 2. Missing values before imputation for the early prediction of discharge destination.**

| **Variables** | **Missing values** | **Total number** |
| --- | --- | --- |
| discharge_location | 0 (0.00%) | 1485 |
| sum_diagnosis | 7 (0.47%) | 1485 |
| ethnicity | 117 (7.88%) | 1485 |
| age | 7 (0.47%) | 1485 |
| gender | 1 (0.07%) | 1485 |
| careunit | 293 (19.7%) | 1485 |
| WBC | 37 (2.49%) | 1485 |
| HR | 316 (21.3%) | 1485 |
| respiration | 316 (21.3%) | 1485 |
| Arterial.BP..Diastolic. | 321 (21.6%) | 1485 |
| Arterial.BP..Systolic. | 324 (21.8%) | 1485 |
| Arterial.BP..mean. | 319 (21.5%) | 1485 |
| Potassium | 42 (2.83%) | 1485 |
| Sodium | 43 (2.90%) | 1485 |
| PO2 | 715 (48.1%) | 1485 |
| PT | 282 (19.0%) | 1485 |
| PTT | 386 (26.0%) | 1485 |
| Glucose.Blood | 40 (2.69%) | 1485 |
| Hemoglobin | 35 (2.36%) | 1485 |
| RDW | 62 (4.18%) | 1485 |
| BUN | 43 (2.90%) | 1485 |
| platelets | 42 (2.83%) | 1485 |
| creatinine | 41 (2.76%) | 1485 |
| bicarbonate | 69 (4.65%) | 1485 |
| hematocrit | 34 (2.29%) | 1485 |
| lactate | 665 (44.8%) | 1485 |
| INR | 282 (19.0%) | 1485 |
| BE | 751 (50.6%) | 1485 |
| Basophils | 502 (33.8%) | 1485 |
| Ca | 79 (5.32%) | 1485 |
| Chloride | 46 (3.10%) | 1485 |
| Free.Ca | 848 (57.1%) | 1485 |
| Phosphate | 345 (23.2%) | 1485 |
| Eosinophils | 490 (33.0%) | 1485 |
| Lymphocytes | 492 (33.1%) | 1485 |
| Magnesium | 230 (15.5%) | 1485 |
| Monocytes | 494 (33.3%) | 1485 |
| PCO2 | 717 (48.3%) | 1485 |
| RBC | 41 (2.76%) | 1485 |
| MCH | 86 (5.79%) | 1485 |
| MCHC | 377 (25.4%) | 1485 |
| MCV | 55 (3.70%) | 1485 |
| PH | 699 (47.1%) | 1485 |
| Anion.Gap | 175 (11.8%) | 1485 |
| CO2.Blood | 882 (59.4%) | 1485 |
| Specific.Gravity | 580 (39.1%) | 1485 |
| GCS.total | 319 (21.5%) | 1485 |
| source.of.admission | 174 (11.7%) | 1485 |
| Paralysis | 0 (0.00%) | 1485 |
| Mechanical.ventilation | 0 (0.00%) | 1485 |
| Weight | 400 (26.9%) | 1485 |
| Height | 558 (37.6%) | 1485 |
| BMI | 600 (40.4%) | 1485 |
| Fio2 | 808 (54.4%) | 1485 |
| Total.bilirubin | 658 (44.3%) | 1485 |
| Albumin | 693 (46.7%) | 1485 |
| Morphine.Sulfate | 0 (0.00%) | 1485 |
| cefazolin | 0 (0.00%) | 1485 |
| Kcl | 0 (0.00%) | 1485 |
| glucocorticoid | 0 (0.00%) | 1485 |
| Dopamine | 0 (0.00%) | 1485 |
| Dobutamine | 0 (0.00%) | 1485 |
| Epinephrine | 0 (0.00%) | 1485 |
| Norepinephrine | 0 (0.00%) | 1485 |

**Supplementary Table 3. Missing values before imputation for the recent prediction of discharge destination.**

| **Variables** | **Missing values** | **Total number** |
| --- | --- | --- |
| discharge_destination | 0 (0.00%) | 1485 |
| los_hospital | 0 (0.00%) | 1485 |
| los_icustays | 297 (20.0%) | 1485 |
| sum_diagnosis | 7 (0.47%) | 1485 |
| ethnicity | 117 (7.88%) | 1485 |
| age | 7 (0.47%) | 1485 |
| gender | 1 (0.07%) | 1485 |
| careunit | 291 (19.6%) | 1485 |
| WBC | 40 (2.69%) | 1485 |
| HR | 312 (21.0%) | 1485 |
| respiration | 311 (20.9%) | 1485 |
| Arterial.BP..Diastolic. | 320 (21.5%) | 1485 |
| Arterial.BP..Systolic. | 321 (21.6%) | 1485 |
| Arterial.BP..mean. | 317 (21.3%) | 1485 |
| Potassium | 48 (3.23%) | 1485 |
| Sodium | 48 (3.23%) | 1485 |
| PO2 | 728 (49.0%) | 1485 |
| PT | 320 (21.5%) | 1485 |
| PTT | 418 (28.1%) | 1485 |
| Glucose.Blood | 39 (2.63%) | 1485 |
| Hemoglobin | 40 (2.69%) | 1485 |
| RDW | 66 (4.44%) | 1485 |
| BUN | 50 (3.37%) | 1485 |
| platelets | 47 (3.16%) | 1485 |
| creatinine | 49 (3.30%) | 1485 |
| bicarbonate | 73 (4.92%) | 1485 |
| hematocrit | 39 (2.63%) | 1485 |
| lactate | 703 (47.3%) | 1485 |
| INR | 320 (21.5%) | 1485 |
| BE | 763 (51.4%) | 1485 |
| Basophils | 518 (34.9%) | 1485 |
| Ca | 83 (5.59%) | 1485 |
| Chloride | 51 (3.43%) | 1485 |
| Free.Ca | 860 (57.9%) | 1485 |
| Phosphate | 344 (23.2%) | 1485 |
| Eosinophils | 503 (33.9%) | 1485 |
| Lymphocytes | 508 (34.2%) | 1485 |
| Magnesium | 231 (15.6%) | 1485 |
| Monocytes | 510 (34.3%) | 1485 |
| PCO2 | 732 (49.3%) | 1485 |
| RBC | 45 (3.03%) | 1485 |
| MCH | 91 (6.13%) | 1485 |
| MCHC | 382 (25.7%) | 1485 |
| MCV | 60 (4.04%) | 1485 |
| PH | 714 (48.1%) | 1485 |
| Anion.Gap | 186 (12.5%) | 1485 |
| CO2.Blood | 886 (59.7%) | 1485 |
| Specific.Gravity | 587 (39.5%) | 1485 |
| GCS.total | 367 (24.7%) | 1485 |
| source.of.admission | 174 (11.7%) | 1485 |
| Los.before.ICU.admission | 291 (19.6%) | 1485 |
| Los.after.ICU.discharge | 291 (19.6%) | 1485 |
| Paralysis | 0 (0.00%) | 1485 |
| Mechanical.ventilation | 0 (0.00%) | 1485 |
| Weight | 694 (46.7%) | 1485 |
| Height | 558 (37.6%) | 1485 |
| BMI | 898 (60.5%) | 1485 |
| Fio2 | 822 (55.4%) | 1485 |
| Total.bilirubin | 682 (45.9%) | 1485 |
| Albumin | 714 (48.1%) | 1485 |
| Morphine.Sulfate | 0 (0.00%) | 1485 |
| cefazolin | 0 (0.00%) | 1485 |
| Kcl | 0 (0.00%) | 1485 |
| glucocorticoid | 0 (0.00%) | 1485 |
| Dopamine | 0 (0.00%) | 1485 |
| Dobutamine | 0 (0.00%) | 1485 |
| Epinephrine | 0 (0.00%) | 1485 |
| Norepinephrine | 0 (0.00%) | 1485 |

**Supplementary Table 4. Characteristics of patients for MIMIC-III-IV and EICU.**

|  | **level** | **Early prediction** | | | **Recent prediction** | | |
| --- | --- | --- | --- | --- | --- | --- | --- |
|  |  | **MIMIC-III-IV (n=800)** | **EICU (n=685)** | **p** | **MIMIC-III-IV (n=800)** | **EICU (n=685)** | **p** |
| discharge_destination (%) | Dead | 78 (9.8) | 73 (10.7) | <0.001 | 78 (9.8) | 73 (10.7) | <0.001 |
|  | Further medical care | 500 (62.5) | 302 (44.1) |  | 500 (62.5) | 302 (44.1) |  |
|  | Home | 222 (27.8) | 310 (45.3) |  | 222 (27.8) | 310 (45.3) |  |
| sum_diagnosis (mean (SD)) |  | 15.66 (8.47) | 6.35 (6.50) | <0.001 | 15.66 (8.47) | 6.31 (6.42) | <0.001 |
| ethnicity (%) | African American | 95 (11.9) | 109 (15.9) | 0.235 | 89 (11.1) | 106 (15.5) | 0.146 |
|  | Asian | 21 (2.6) | 17 (2.5) |  | 21 (2.6) | 16 (2.3) |  |
|  | Hispanic | 47 (5.9) | 33 (4.8) |  | 48 (6.0) | 33 (4.8) |  |
|  | Other | 40 (5.0) | 33 (4.8) |  | 38 (4.8) | 33 (4.8) |  |
|  | White | 597 (74.6) | 493 (72.0) |  | 604 (75.5) | 497 (72.6) |  |
| age (years) (mean (SD)) |  | 56.47 (18.97) | 55.64 (19.05) | 0.403 | 56.38 (18.93) | 55.64 (19.05) | 0.457 |
| gender (%) | Female | 216 (27.0) | 208 (30.4) | 0.17 | 216 (27.0) | 207 (30.2) | 0.189 |
|  | Male | 584 (73.0) | 477 (69.6) |  | 584 (73.0) | 478 (69.8) |  |
| Weight (Kg) (mean (SD)) |  | 84.83 (27.90) | 82.56 (23.13) | 0.091 | 85.63 (28.24) | 80.68 (25.24) | <0.001 |
| Height (cm) (mean (SD)) |  | 173.21 (11.49) | 172.28 (14.09) | 0.161 | 172.49 (11.74) | 172.22 (14.08) | 0.691 |
| BMI (kg/㎡) (mean (SD)) |  | 28.02 (6.80) | 27.89 (8.27) | 0.734 | 28.37 (6.99) | 27.15 (7.87) | 0.002 |
| careunit (%) | CCU | 96 (12.0) | 51 (7.4) | 0.013 | 87 (10.9) | 50 (7.3) | 0.038 |
|  | MICU/SICU | 555 (69.4) | 495 (72.3) |  | 547 (68.4) | 501 (73.1) |  |
|  | NICU | 149 (18.6) | 139 (20.3) |  | 166 (20.8) | 134 (19.6) |  |
| source.of.admission (%) | Emergency | 575 (71.9) | 480 (70.1) | <0.001 | 574 (71.8) | 479 (69.9) | <0.001 |
|  | General care area/OR | 92 (11.5) | 170 (24.8) |  | 93 (11.6) | 169 (24.7) |  |
|  | Transfer | 133 (16.6) | 35 (5.1) |  | 133 (16.6) | 37 (5.4) |  |
| Paralysis (%) | No | 562 (70.2) | 385 (56.2) | <0.001 | 562 (70.2) | 384 (56.1) | <0.001 |
|  | Yes | 238 (29.8) | 300 (43.8) |  | 238 (29.8) | 301 (43.9) |  |
| Mechanical.ventilation (%) | No | 447 (55.9) | 422 (61.6) | 0.029 | 447 (55.9) | 414 (60.4) | 0.085 |
|  | Yes | 353 (44.1) | 263 (38.4) |  | 353 (44.1) | 271 (39.6) |  |
| GCS.total (mean (SD)) |  | 12.13 (4.22) | 12.38 (4.14) | 0.252 | 13.60 (3.06) | 12.33 (4.21) | <0.001 |
| HR (bpm) (mean (SD)) |  | 82.54 (20.23) | 87.09 (21.58) | <0.001 | 75.00 (25.87) | 78.03 (26.82) | 0.027 |
| respiration (bpm) (mean (SD)) |  | 18.00 (5.75) | 17.36 (9.30) | 0.107 | 17.78 (7.59) | 15.90 (9.42) | <0.001 |
| Arterial.BP..Diastolic. (mmHg) (mean (SD)) | | 68.51 (17.32) | 70.89 (19.77) | 0.013 | 62.32 (18.73) | 69.63 (16.39) | <0.001 |
| Arterial.BP..Systolic. (mmHg) (mean (SD)) | | 131.19 (25.89) | 123.31 (28.10) | <0.001 | 119.93 (31.44) | 125.29 (23.28) | <0.001 |
| Arterial.BP..mean. (mmHg) (mean (SD)) | | 85.78 (19.83) | 85.12 (21.34) | 0.54 | 77.28 (21.71) | 84.87 (17.81) | <0.001 |
| Potassium (mmol/L) (mean (SD)) | | 4.10 (0.65) | 4.05 (0.63) | 0.142 | 4.15 (0.49) | 4.05 (0.49) | <0.001 |
| Sodium (mmol/L) (mean (SD)) |  | 138.83 (4.37) | 137.63 (4.72) | <0.001 | 138.65 (4.00) | 138.51 (4.37) | 0.522 |
| PO2 (mmHg) (mean (SD)) |  | 168.76 (106.74) | 143.97 (96.64) | <0.001 | 129.46 (68.60) | 112.60 (64.89) | <0.001 |
| PT (seconds) (mean (SD)) |  | 13.89 (5.35) | 14.51 (5.53) | 0.027 | 13.89 (6.21) | 14.39 (4.12) | 0.073 |
| PTT (seconds) (mean (SD)) |  | 30.19 (14.14) | 31.53 (14.28) | 0.069 | 31.28 (12.60) | 31.99 (12.54) | 0.278 |
| Glucose.Blood (mg/dL) (mean (SD)) | | 139.15 (120.01) | 135.79 (59.51) | 0.506 | 123.24 (39.51) | 126.61 (49.91) | 0.147 |
| Hemoglobin (g/dL) (mean (SD)) | | 11.96 (2.37) | 12.22 (2.26) | 0.032 | 10.64 (1.91) | 10.83 (2.04) | 0.063 |
| RDW (%) (mean (SD)) |  | 13.95 (1.88) | 14.74 (2.26) | <0.001 | 14.49 (1.90) | 15.17 (2.53) | <0.001 |
| BUN (mg/dL) (mean (SD)) |  | 18.62 (13.33) | 20.70 (16.03) | 0.006 | 19.51 (15.61) | 18.21 (13.58) | 0.088 |
| WBC (K/uL) (mean (SD)) |  | 11.07 (5.86) | 12.27 (6.91) | <0.001 | 9.82 (5.00) | 9.85 (5.18) | 0.911 |
| Platelets (K/uL) (mean (SD)) |  | 228.05 (102.65) | 244.48 (115.28) | 0.004 | 282.89 (152.81) | 252.43 (123.05) | <0.001 |
| creatinine (mg/dL) (mean (SD)) | | 0.99 (0.63) | 1.10 (1.06) | 0.009 | 0.83 (0.58) | 0.88 (0.81) | 0.235 |
| bicarbonate (mmol/L) (mean (SD)) | | 23.97 (4.00) | 24.29 (4.62) | 0.152 | 26.77 (3.79) | 26.62 (4.16) | 0.461 |
| hematocrit (%) (mean (SD)) |  | 35.63 (6.69) | 36.79 (6.24) | 0.001 | 32.13 (5.30) | 32.90 (5.76) | 0.008 |
| lactate (mmol/L) (mean (SD)) |  | 1.97 (1.28) | 2.05 (1.70) | 0.268 | 1.66 (1.27) | 1.61 (1.72) | 0.508 |
| INR (mean (SD)) |  | 1.28 (0.66) | 1.25 (0.54) | 0.357 | 1.26 (0.62) | 1.23 (0.42) | 0.291 |
| BE (mmol/L) (mean (SD)) |  | -0.93 (3.72) | -0.64 (6.55) | 0.287 | 1.54 (4.16) | 1.03 (5.37) | 0.041 |
| Basophils (%) (mean (SD)) |  | 0.35 (0.37) | 0.30 (0.40) | 0.027 | 0.38 (0.41) | 0.34 (0.44) | 0.088 |
| Ca (mg/dL) (mean (SD)) |  | 8.48 (0.79) | 8.64 (0.82) | <0.001 | 8.65 (0.71) | 8.59 (0.66) | 0.112 |
| Chloride (mmol/L) (mean (SD)) | | 103.93 (5.26) | 103.08 (5.82) | 0.003 | 102.19 (4.78) | 103.63 (5.29) | <0.001 |
| Free.Ca (mmol/L) (mean (SD)) |  | 1.10 (0.16) | 0.76 (0.59) | <0.001 | 1.12 (0.09) | 0.78 (0.47) | <0.001 |
| Phosphate (mg/dL) (mean (SD)) | | 3.48 (1.06) | 3.20 (1.09) | <0.001 | 3.53 (1.04) | 3.30 (0.98) | <0.001 |
| Eosinophils (%) (mean (SD)) |  | 1.37 (1.72) | 1.34 (1.86) | 0.737 | 1.68 (1.94) | 2.29 (2.63) | <0.001 |
| Lymphocytes (%) (mean (SD)) |  | 13.74 (9.95) | 16.55 (11.42) | <0.001 | 14.54 (9.65) | 17.64 (11.64) | <0.001 |
| Magnesium (mg/dL) (mean (SD)) | | 1.83 (0.32) | 1.85 (0.34) | 0.413 | 2.03 (0.27) | 1.98 (0.31) | 0.004 |
| Monocytes (%) (mean (SD)) |  | 5.47 (3.05) | 7.17 (3.25) | <0.001 | 5.60 (2.88) | 8.39 (4.33) | <0.001 |
| PCO2 (mmHg) (mean (SD)) |  | 42.02 (10.50) | 43.44 (11.98) | 0.015 | 42.79 (9.71) | 39.88 (8.76) | <0.001 |
| RBC (m/uL) (mean (SD)) |  | 3.96 (0.78) | 4.11 (0.73) | <0.001 | 3.56 (0.65) | 3.68 (0.67) | <0.001 |
| MCH (pg) (mean (SD)) |  | 30.15 (3.39) | 29.66 (2.75) | 0.002 | 30.08 (2.44) | 29.54 (2.49) | <0.001 |
| MCHC (%) (mean (SD)) |  | 33.46 (2.00) | 33.14 (1.40) | <0.001 | 33.09 (1.64) | 32.88 (1.33) | 0.009 |
| MCV (fL) (mean (SD)) |  | 90.08 (9.22) | 89.46 (6.85) | 0.148 | 90.98 (6.40) | 89.77 (6.21) | <0.001 |
| PH (mean (SD)) |  | 7.37 (0.08) | 7.37 (0.11) | 0.712 | 7.40 (0.07) | 7.42 (0.08) | 0.003 |
| Anion.Gap (mmol/L) (mean (SD)) | | 14.51 (3.36) | 11.24 (4.40) | <0.001 | 13.12 (3.11) | 9.30 (4.00) | <0.001 |
| CO2.Blood (mmol/L) (mean (SD)) | | 25.09 (4.11) | 27.44 (8.51) | <0.001 | 27.63 (4.78) | 28.75 (9.30) | 0.003 |
| Specific.Gravity (mean (SD)) |  | 1.02 (0.01) | 1.02 (0.01) | 0.102 | 1.02 (0.01) | 1.02 (0.01) | <0.001 |
| Fio2 (%) (mean (SD)) |  | 64.62 (25.63) | 45.48 (31.71) | <0.001 | 49.42 (19.92) | 37.25 (23.97) | <0.001 |
| Total.bilirubin (mg/dL) (mean (SD)) | | 0.80 (1.12) | 0.63 (0.69) | 0.001 | 0.82 (2.09) | 0.75 (1.36) | 0.454 |
| Albumin (g/dL) (mean (SD)) |  | 3.28 (0.68) | 3.18 (0.73) | 0.008 | 3.33 (0.70) | 2.96 (0.72) | <0.001 |
| Morphine.Sulfate (%) | No | 408 (51.0) | 609 (88.9) | <0.001 | 408 (51.0) | 612 (89.3) | <0.001 |
|  | Yes | 392 (49.0) | 76 (11.1) |  | 392 (49.0) | 73 (10.7) |  |
| cefazolin (%) | No | 455 (56.9) | 616 (89.9) | <0.001 | 455 (56.9) | 624 (91.1) | <0.001 |
|  | Yes | 345 (43.1) | 69 (10.1) |  | 345 (43.1) | 61 (8.9) |  |
| Kcl (%) | No | 258 (32.2) | 459 (67.0) | <0.001 | 258 (32.2) | 461 (67.3) | <0.001 |
|  | Yes | 542 (67.8) | 226 (33.0) |  | 542 (67.8) | 224 (32.7) |  |
| glucocorticoid (%) | No | 515 (64.4) | 593 (86.6) | <0.001 | 515 (64.4) | 600 (87.6) | <0.001 |
|  | Yes | 285 (35.6) | 92 (13.4) |  | 285 (35.6) | 85 (12.4) |  |
| Dopamine (%) | No | 770 (96.2) | 670 (97.8) | 0.11 | 770 (96.2) | 671 (98.0) | 0.075 |
|  | Yes | 30 (3.8) | 15 (2.2) |  | 30 (3.8) | 14 (2.0) |  |
| Dobutamine (%) | No | 794 (99.2) | 683 (99.7) | 0.397 | 794 (99.2) | 683 (99.7) | 0.397 |
|  | Yes | 6 (0.8) | 2 (0.3) |  | 6 (0.8) | 2 (0.3) |  |
| Epinephrine (%) | No | 768 (96.0) | 672 (98.1) | 0.028 | 768 (96.0) | 670 (97.8) | 0.066 |
|  | Yes | 32 (4.0) | 13 (1.9) |  | 32 (4.0) | 15 (2.2) |  |
| Norepinephrine (%) | No | 661 (82.6) | 583 (85.1) | 0.221 | 661 (82.6) | 590 (86.1) | 0.076 |
|  | Yes | 139 (17.4) | 102 (14.9) |  | 139 (17.4) | 95 (13.9) |  |
| los_hospital (days) (mean (SD)) | |  |  |  | 12.13 (11.68) | 11.61 (14.94) | 0.454 |
| los_icustays (days) (mean (SD)) | |  |  |  | 6.48 (8.13) | 5.13 (9.42) | 0.003 |
| Los.before.ICU.admission (days) (mean (SD)) | | |  |  | 0.96 (2.55) | 0.77 (2.83) | 0.176 |
| Los.after.ICU.discharge (days) (mean (SD)) | | |  |  | 4.22 (6.67) | 4.71 (8.33) | 0.209 |

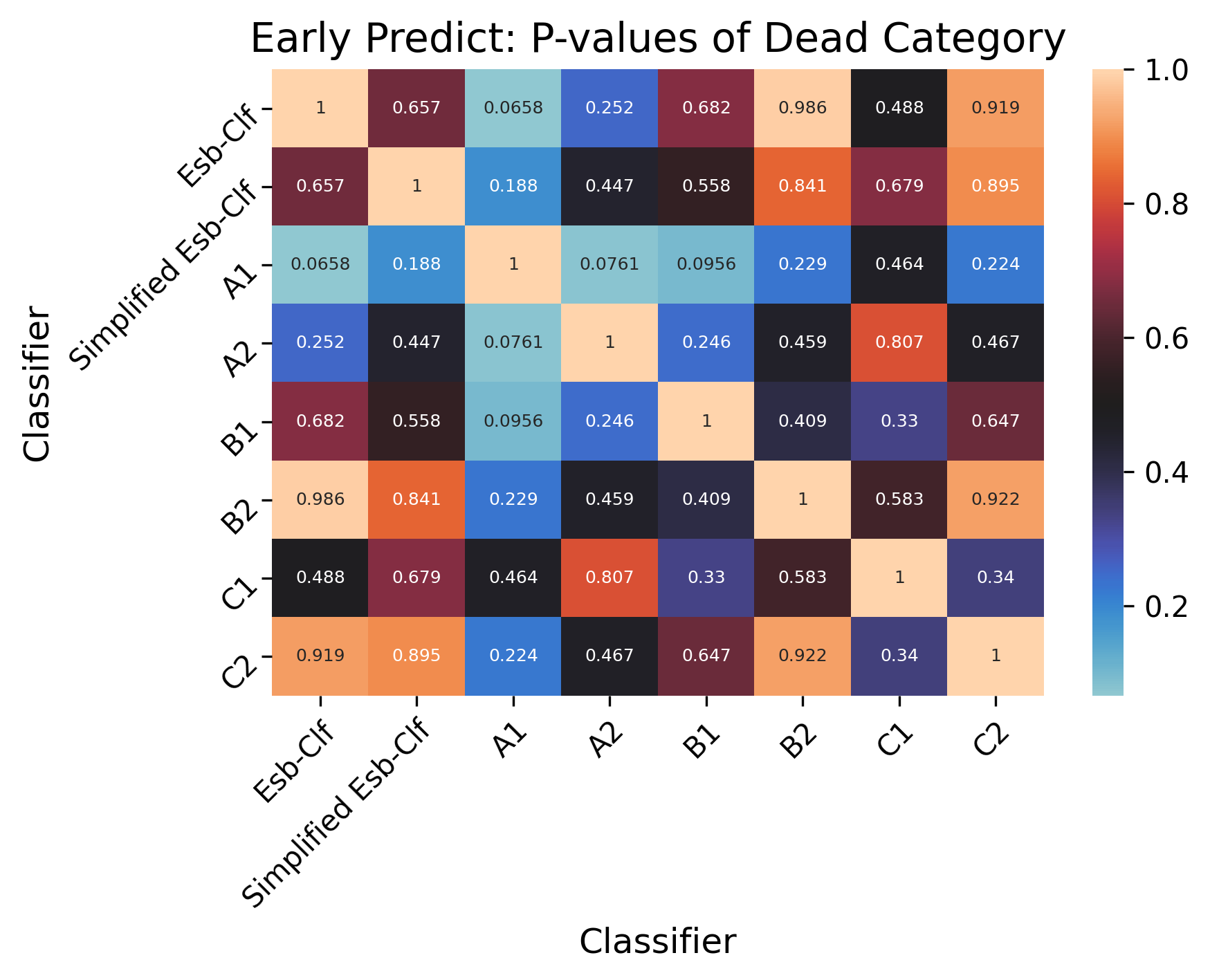

**Supplementary Fig 1. Delong test of the early prediction of dead.**

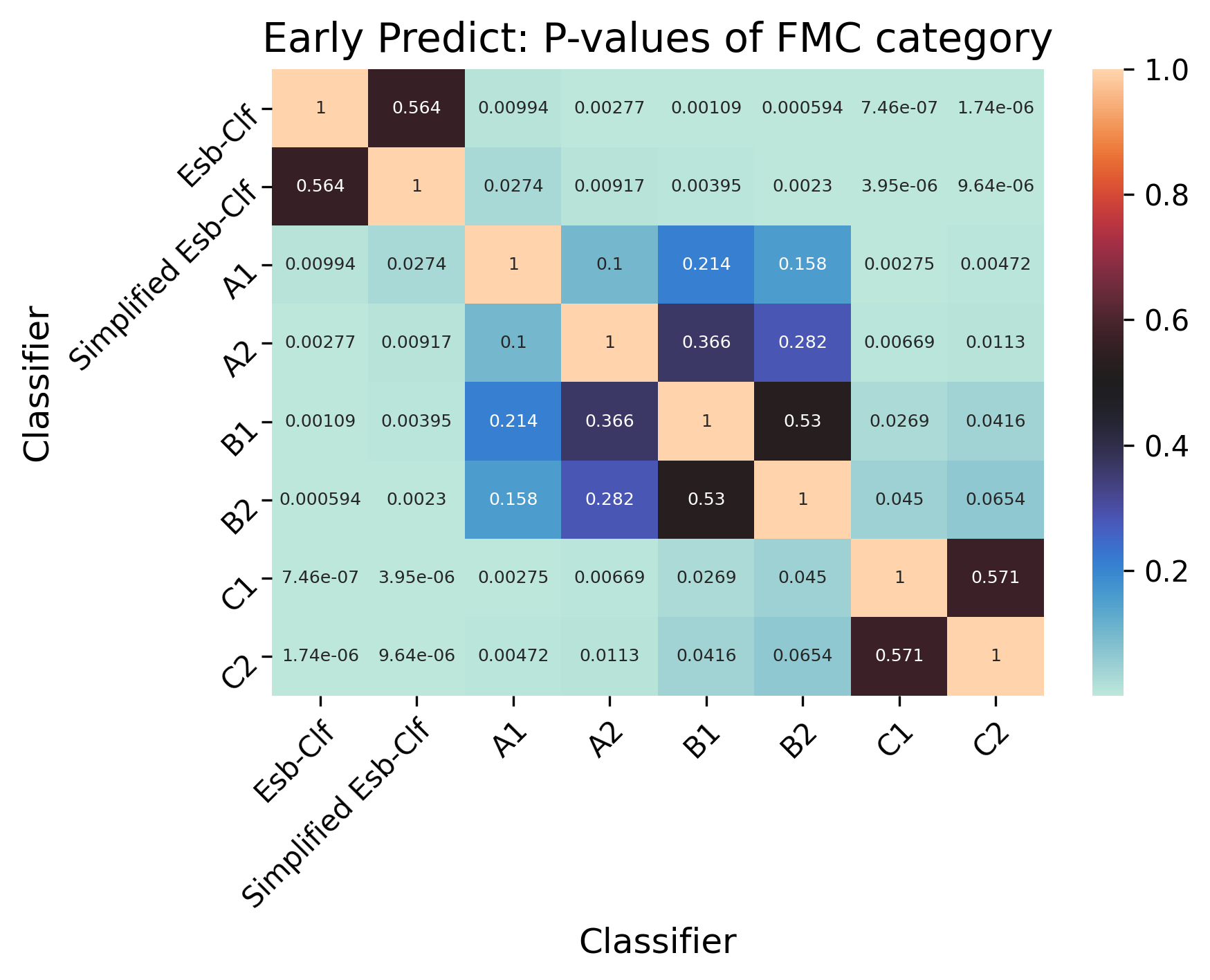

**Supplementary Fig 2. Delong test of the early prediction of further medical care (FMC).**

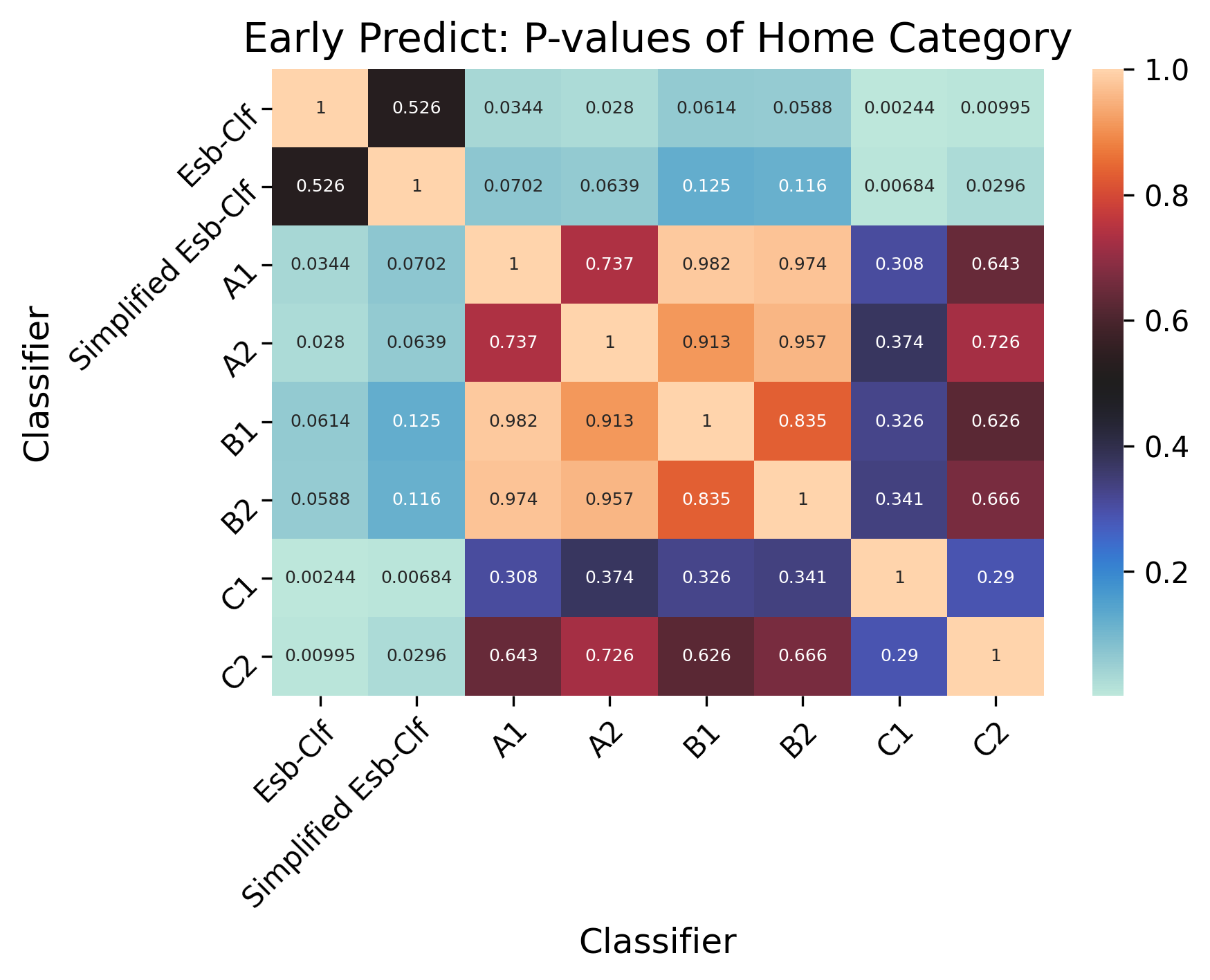

**Supplementary Fig 3. Delong test of the early prediction of home.**

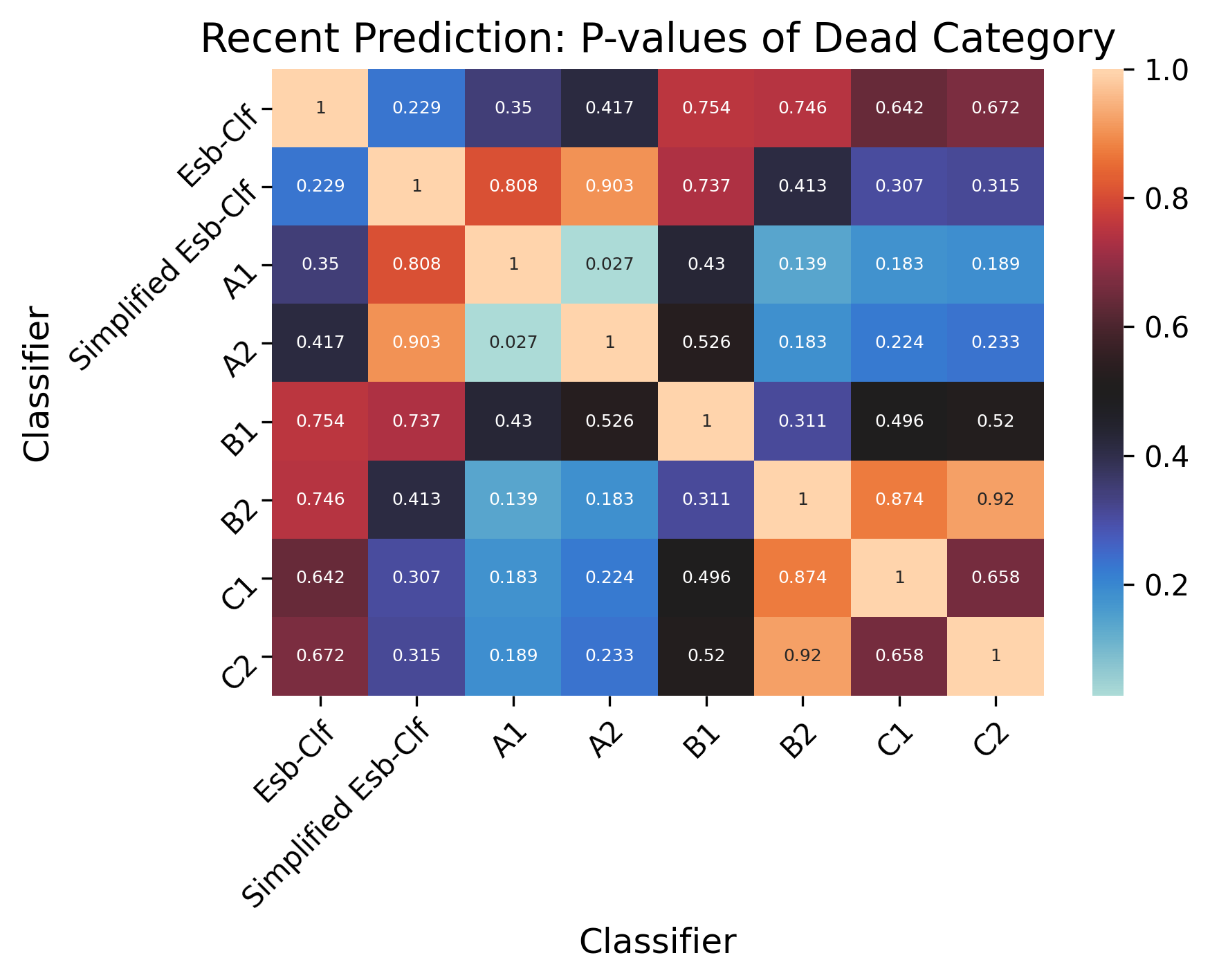

**Supplementary Fig 4. Delong test of the recent prediction of dead.**

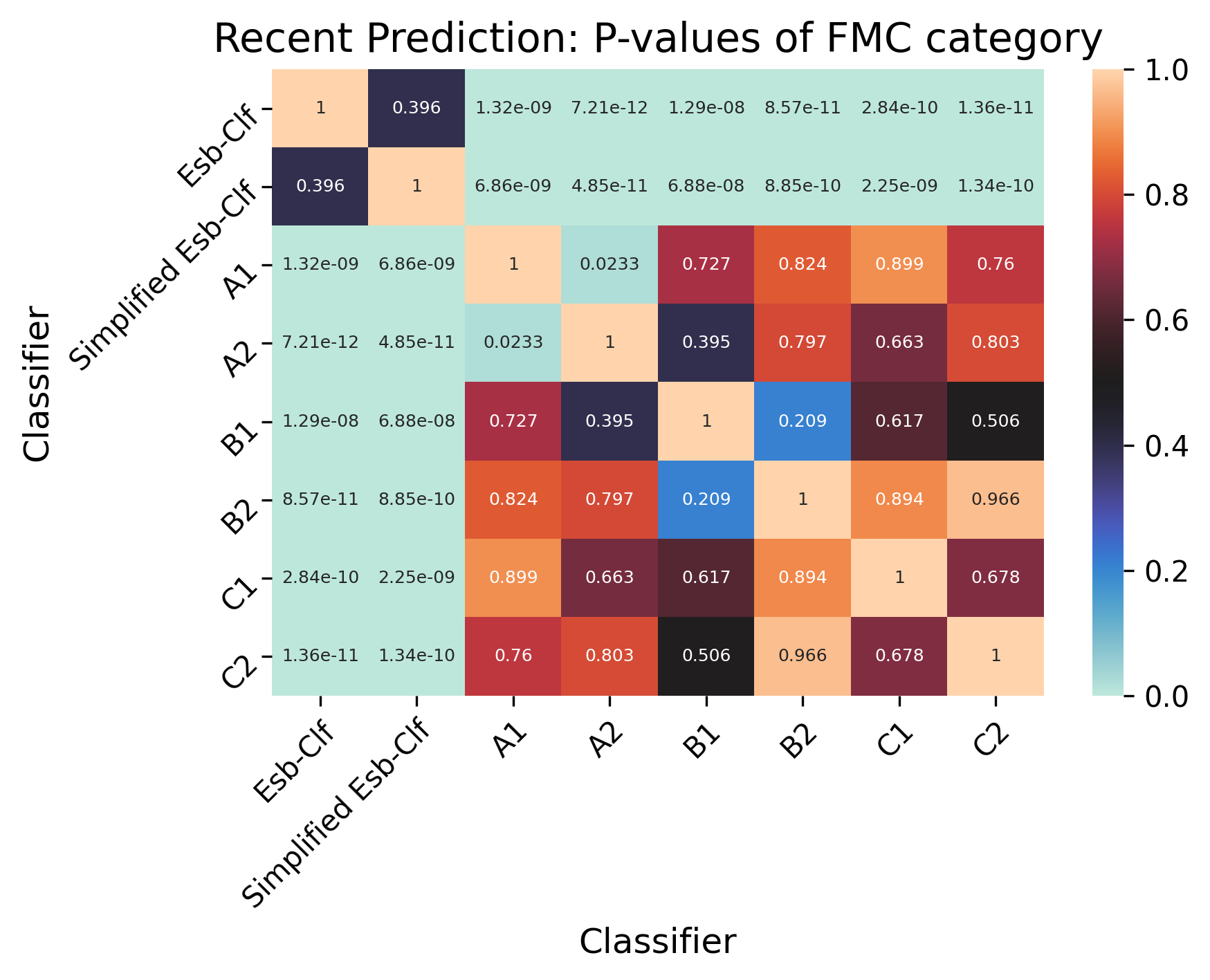

**Supplementary Fig 5. Delong test of the recent prediction of further medical care (FMC).**

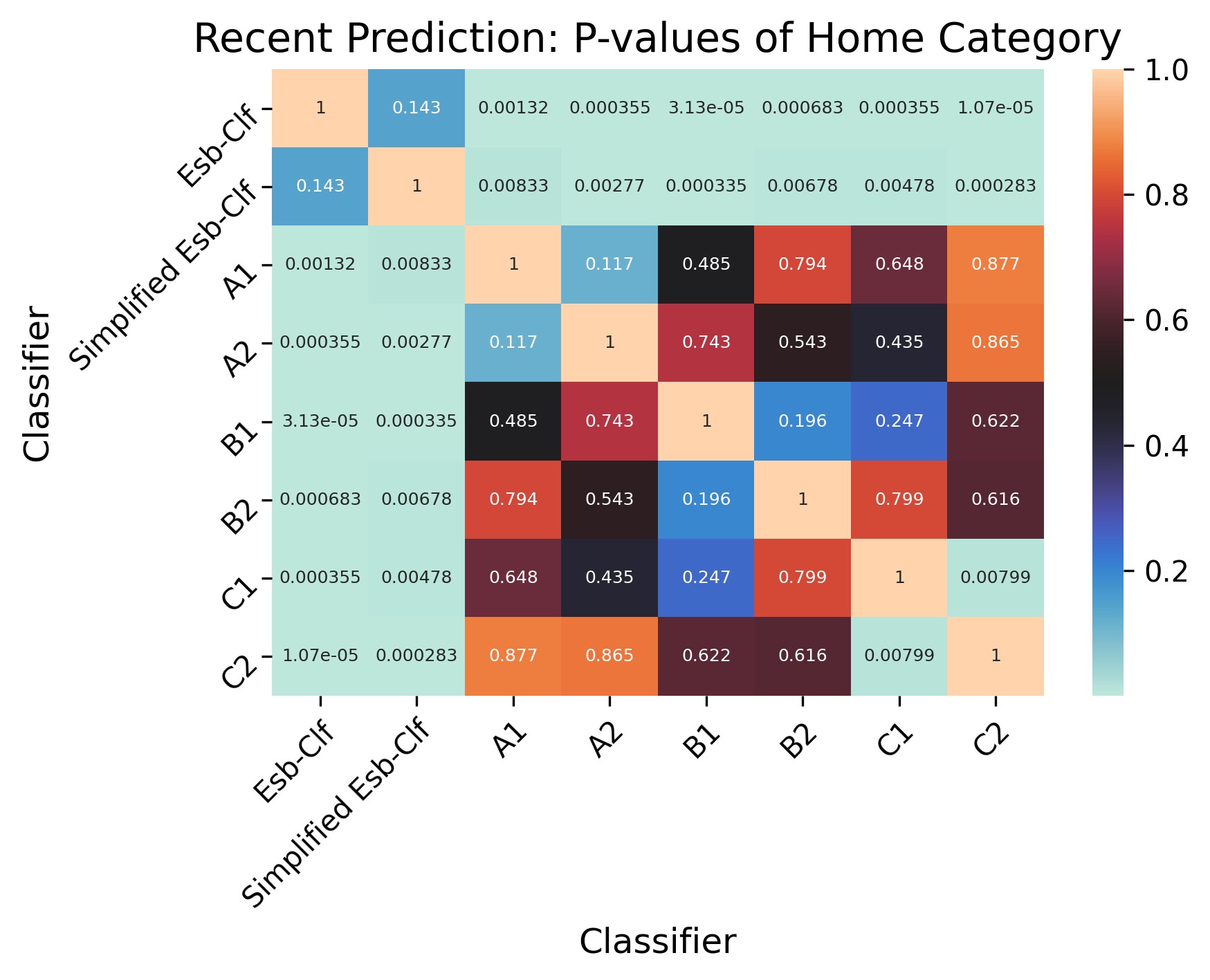

**Supplementary Fig 6. Delong test of the recent prediction of home.**
