## Supplementary material for "Discharge prediction of critical patients with spinal cord injury: a machine learning study with 1485 cases": coi_disclosure

**ICMJE DISCLOSURE FORM**

**Date:_ 2021/06/26____________________________________________________________________**

**Manuscript number (if known):__________________________________________________________________**

**In the interest of transparency, we ask you to disclose all relationships/activities/interests listed below that are**

**related to the content of your manuscript. “Related” means any relation with for-profit or not-for-profit third**

**parties whose interests may be affected by the content of the manuscript. Disclosure represents a commitment**

**to transparency and does not necessarily indicate a bias. If you are in doubt about whether to list a relationship/activity/interest, it is preferable that you do so.**

**The following questions apply to the** **author’s relationships/activities/interests as they relate to the current**

**manuscript only.**

**The author’s relationships/activities/interests should be defined broadly. For example, if your manuscript pertains**

**to the epidemiology of hypertension, you should declare all relationships with manufacturers of antihypertensive medication, even if that medication is not mentioned in the manuscript.**

**In item #1 below, report all support for the work reported in this manuscript without time limit. For all other items,**

**the time frame for disclosure is the past 36 months.**

|  |  | **Name all entities with whom you have this relationship or indicate none (add rows as needed)** | | **Specifications/Comments**  **(e.g., if payments were made to you or to your institution)** |
| --- | --- | --- | --- | --- |
| **Time frame: Since the initial planning of the work** | | | | |
| 1 | All support for the present manuscript (e.g., funding, provision of study materials, medical writing, article processing charges, etc.)  **No time limit for this item.** | ____ None | |  |
| **Time frame: past 36 months** | | | | |
| 2 | Grants or contracts from any entity (if not indicated in item #1 above). | ____ None |  | |
| 3 | Royalties or licenses | ____ None |  | |
| 4 | Consulting fees | ____ None |  | |
| 5 | Payment or honoraria for lectures, presentations, speakers bureaus, manuscript writing or educational events | ____ None |  | |
| 6 | Payment for expert testimony | ____ None |  | |
| 7 | Support for attending meetings and/or travel | ____ None |  | |
| 8 | Patents planned, issued or pending | ____ None |  | |
| 9 | Participation on a Data  Safety Monitoring Board or Advisory Board | ____ None |  | |
| 10 | Leadership or fiduciary role in other board, society, committee or advocacy group, paid or unpaid | ____ None |  | |
| 11 | Stock or stock options | ____ None |  | |
| 12 | Receipt of equipment, materials, drugs, medical writing, gifts or other services | ____ None |  | |
| 13 | Other financial or non-financial interests | ____ None |  | |

**Please place an “X” next to the following statement to indicate your agreement:**

**X __ I certify that I have answered every question and have not altered the wording of any of the questions on this**

**form.**
